# A Controlled Human Malaria Infection model for relapsing *Plasmodium vivax*

**DOI:** 10.64898/2026.06.18.26355868

**Authors:** Andrew D.S. Duncan, Tessa J.M. Geraedts, Denise Manlutac, Ellie C. Baker, Jennifer K. van Heerden, Thomas W. Roberts, Cassandra A. Rigby, Jo Salkeld, Mimi M. Hou, Clare Megson, Kavyashri Kashojhala, Jodie Boyle, Chidimma Nwankwo, Barnabas G. Williams, Kirsty McHugh, Zi Wei Teo, Ana S. Rodrigues, Camilla A. Gladstone, Ebrima Comma, Yama F. Mujadidi, Hannah Robinson, Emma Plested, Matthew B.B. McCall, Teun Bousema, Markus Gmeiner, Jeroen Bok, Wouter Graumans, Geert-Jan van Gemert, Rachel E. Cowan, Amy Boyd, Phebe Ekregbesi, Nelly Owino, Jee-Sun Cho, Fay L. Nugent, Carolyn M. Nielsen, Benjamin Mordmüller, Francesca R. Donnellan, Sarah E. Silk, Simon J. Draper, Angela M. Minassian

## Abstract

**Background:** *Plasmodium vivax* malaria relapses are a major source of morbidity and onward transmission of infection. The underlying mechanisms are poorly understood and current therapies sub-optimal. We examined the safety and feasibility of a controlled human malaria infection (CHMI) model for relapsing *P. vivax*.

**Methods:** We conducted an open-label, proof-of-concept, CHMI study of relapsing *P. vivax*. Healthy, malaria-naïve, Duffy-positive adults aged 18–45 years with extensive CYP2D6 metaboliser phenotype and normal blood glucose-6-phosphate dehydrogenase (G6PD) levels were recruited in Oxford, UK. Mosquito-bite CHMI was performed in Nijmegen, The Netherlands, using *Anopheles stephensi* mosquitoes infected with PvW1, a clonal isolate of *P. vivax* from Thailand. All follow-up visits were conducted in Oxford, UK. Primary *P. vivax* infections (qPCR > 500 genome copies/mL) were treated with artemether-lumefantrine (80mg/480mg at 8, 24, 36, 48 and 60 hours). From Day 28 following CHMI, participants attended a fortnightly clinic for clinical review and qPCR blood sampling, with additional assessments performed for any reported symptoms. *P. vivax* relapse infections (qPCR > 500 genome copies/mL) were treated with artemether-lumefantrine as per primary infection. Definitive anti-malarial treatment with atovaquone-proguanil (1000mg/400mg once daily for three days) and primaquine (0·5 mg/kg/day for 14 days) was administered six months following CHMI, regardless of parasitaemia or symptoms. The primary objective was to assess the safety, feasibility and frequency of relapsing *P. vivax* after CHMI. Remote follow-up (5 years) is ongoing. The study is registered with ISRCTN registry (ISRCTN48625883).

**Findings:** 20 participants were screened for eligibility from 21 January 2025. Five participants (median age 22 years) underwent CHMI (five infected mosquitoes per participant) on 15 April 2025. All participants developed primary *P. vivax* infection and experienced at least one relapse infection. Two participants experienced a second relapse. Overall incidence rate was 3·6 relapse infections per person-year. Solicited adverse events were mild or moderate and there were no serious adverse events. Definitive anti-malarial treatment was administered to all participants. One participant experienced primaquine-induced methaemoglobinaemia, resolving with early discontinuation of treatment (total dose 5·3 mg/kg). To date, more than six months after primaquine treatment, no further relapses have been recorded.

**Interpretation:** CHMI of relapsing *P. vivax* is safe and feasible, allowing exploration of the mechanisms underlying relapse infections and providing a platform for future anti-relapse efficacy studies.

**Funding:** European Union Horizon Europe programme and UK Research and Innovation (UKRI) via OptiViVax consortium; UK National Institute for Health and Care Research Biomedical Research Centre: Oxford; and UK Medical Research Council.

**Research in Context:** *Evidence before this study:* Relapse infections account for the majority of cases of *Plasmodium vivax* malaria. There are no licensed vaccines for *P. vivax* and current anti-relapse hypnozoiticidal therapies (primaquine and tafenoquine) have restrictive contraindications, side effects and variable efficacy. A controlled human malaria infection (CHMI) model for relapsing *P. vivax*, using a well-characterised clonal isolate of *P. vivax* administered by mosquito bite, would present an opportunity to improve understanding of hypnozoite immuno-biology and provide a platform for efficacy studies of novel anti-relapse interventions. We searched PubMed on 23 April 2026 for research articles using the terms (“P. vivax” OR “Plasmodium vivax” OR “vivax”) AND (“malaria challenge” OR “Controlled Human Malaria Infection” OR “CHMI” OR “sporozoite challenge”). No date or language filters were applied. The search identified seven published CHMI studies in which *P. vivax* was administered by mosquito bite. All these studies utilised mosquitoes infected from a source patient(s) with naturally-acquired infection, thereby limiting interstudy comparison and creating logistical challenges for reproducibility. Only one study reported relapse infections which occurred following primaquine treatment in two participants with poor and intermediate CYP2D6 metaboliser phenotype respectively. In addition to this PubMed search, historical records were identified which described the deliberate infection of humans with *P. vivax* i) in experiments conducted in the early 20^th^ Century, and ii) as part of malariotherapy for the treatment of “general paralysis of the insane” (neurosyphilis). Although some of these records used well-characterised strains of *P. vivax* (e.g. Madagascar strain) and included relapse infections, these pre-dated modern molecular and serological laboratory techniques. Having previously generated a cryopreserved inoculum of *P. vivax*-infected red blood cells (PvW1) with high-quality genome assembly, we set out to administer this clonal isolate in a CHMI study by mosquito bite, and characterise *P. vivax* relapse infections over a six-month period prior to the administration of primaquine treatment.

*Added value of this study:* This study reports the administration of PvW1 by mosquito bite to five healthy adult participants in the UK. All participants developed primary *P. vivax* infection and experienced at least one relapse infection in a six-month follow-up period. Two participants experienced a second relapse infection prior to the administration of definitive anti-malarial treatment with atovaquone-proguanil and primaquine. Solicited adverse events (foreseeable symptoms of malaria) were mild or moderate. Long-term remote follow-up is ongoing. However, to date, more than six months after primaquine treatment, no further relapse infections have been recorded. To our knowledge, this is the first CHMI study in the modern era to administer a well-characterised clonal isolate of *P. vivax* by mosquito bite. We demonstrate that a CHMI model for relapsing *P. vivax* is safe and feasible.

*Implications of all the available evidence:* The World Health Organisation (WHO) Technical Brief on the Control and Elimination of *Plasmodium vivax* Malaria calls for better understanding of the biology and epidemiology of *P. vivax,* and the development of new interventions and strategies. Specifically, it highlights a need for improved knowledge of the underlying mechanisms of *P. vivax* relapse infections; a single-dose hypnozoiticidal treatment that can be used for all population groups without significant side effects; and development of *P. vivax* vaccines. CHMI studies allow the study of host-pathogen interactions in detail and accelerate the development of vaccines and therapeutic drugs by providing evidence of efficacy early in clinical development. This proof-of-concept study presents an opportunity to explore the mechanisms behind hypnozoite formation and activation in humans, and provides a novel clinical platform for future drug and vaccine efficacy studies.

## Introduction

*Plasmodium vivax* is a major cause of morbidity, and short- and long-term mortality, accounting for 8–18 million cases of malaria every year, predominantly in South and South East Asia, South America and East Africa.^1,2^ In an era where *Plasmodium falciparum* has dominated malaria research and public health control efforts, *P. vivax* is becoming increasingly acknowledged globally and presents a significant challenge in countries striving for malaria elimination.^3^

Relapse infections account for 80–90% of *P. vivax* incident cases.^4,5^ Relapses arise from the reactivation of dormant hypnozoites in the liver.^6,7^ Hypnozoites are formed from sporozoites introduced at the infectious mosquito bite. They can establish an active blood-stage infection weeks to months after the initial inoculation. Relapses occur repeatedly in the absence of adequate treatment, acting as a significant source of symptomatic disease and onward transmission.^3^ Effectively tackling the hypnozoite reservoir of infection could markedly reduce *P. vivax* disease burden, particularly in low-transmission settings.^8^ However, the mechanisms behind hypnozoite formation and reactivation are unknown.

Definitive treatment (radical cure) of an individual with *P. vivax* infection requires hypnozoiticidal treatment with an 8-aminoquinoline, namely primaquine or tafenoquine.^9^ Despite primaquine being used for over 80 years, its mechanism of action is poorly understood although it is proposed to be related to the generation of oxidative metabolites.^10^ Both primaquine and tafenoquine are contraindicated in pregnancy, breast feeding, and glucose-6-phosphate dehydrogenase (G6PD) deficiency due to a risk of inducing haemolysis. The geographical distribution of G6PD deficiency significantly overlaps with that of *P. vivax* malaria.^3^ Primaquine requires a treatment course of 7–14 days which creates difficulties for adherence.^9^ Tafenoquine has the advantage of only requiring a single dose, but its long half-life necessitates more stringent G6PD testing prior to treatment.^9^ The efficacy of primaquine is also variable depending on cytochrome P450 enzyme 2D6 (CYP2D6) polymorphisms.^1,3,11^

In recent years, significant attention has been given to optimising the dose (primaquine and tafenoquine)^12–14^ and duration of treatment (primaquine)^15–17^ to maximise effectiveness of anti-relapse therapy. However, the fact remains that these drugs are sub-optimal and, in endemic areas, approximately 1 in 10 can expect a recurrent *P. vivax* infection (reinfection, recrudescence or relapse) in the six months following the best available regimen.^12,13,17^

Controlled human malaria infection (CHMI) studies, in which healthy participants are experimentally infected with malaria (either by mosquito bite, direct inoculation of cryopreserved sporozoites, or intravenous administration of infected red blood cells), present an opportunity to study host-pathogen interactions, and facilitate the assessment of novel therapies and vaccines.^18^ For *P. falciparum* the sporozoite CHMI model (mosquito bite and direct injection) is well established and was significant in the development of recently deployed vaccines RTS,S/AS01 and R21/Matrix-M.^19,20^ CHMI studies of *P. vivax* are less common in the modern era.^18^ This is partly due to biological characteristics of the species, preventing long-term *in vitro* culture. Consequently, for mosquito-bite CHMI studies, which are most representative of natural infection, the mosquitoes need to be infected directly from a source patient with *P. vivax* infection, typically in an endemic country, invariably resulting in a different strain of *P. vivax* being used in each study.^21^ Blood-stage CHMI studies, using an inoculum of cryopreserved red blood cells infected with an isolate of *P. vivax* administered intravenously, offer more controlled and predictable infection, facilitating interstudy comparison, but do not allow the assessment of pre-erythrocytic interventions or relapse infections.^18,21–24^

A reproducible CHMI model using a well-characterised clonal isolate of *P. vivax* administered by mosquito bite would be a valuable tool in the down selection of *P. vivax* interventions. A CHMI model for relapsing *P. vivax* would present an opportunity to improve understanding of hypnozoite immuno-biology and provide a platform for novel anti-relapse efficacy studies.

Here we present the safety and feasibility of a CHMI model for relapsing *P. vivax* using PvW1, a clonal isolate of *P. vivax* originating from Thailand,^22^ administered by mosquito bite.

## Methods

### Study design and participants

We conducted an open-label, proof-of-concept, experimental medicine CHMI study (called “BIO-006”) of relapsing *P. vivax* at the Centre for Clinical Vaccinology and Tropical Medicine (CCVTM), Oxford, United Kingdom, and Radboud University Medical Center (Radboudumc), Nijmegen, The Netherlands (figure 1). Healthy, malaria-naïve, Duffy blood-group positive adults aged 18–45 years with extensive CYP2D6 metaboliser phenotype and normal blood G6PD levels (5·8–18·8 IU/gHb) were recruited in Oxford. The mosquito-bite CHMI was performed in Nijmegen. All other follow-up visits were conducted in Oxford. All participants provided written informed consent at screening and self-reported sex at birth, gender and ethnicity. The full inclusion and exclusion criteria are listed in the appendix (pp 4–6). The study received ethical approval from UK National Health Service Research Ethics Services (Berkshire Research Ethics Committee Ref 24/SC/0355) and METC Oost-Nederland (Medical Ethics Committee of the Eastern Netherlands Ref 2024-17854). The study was conducted according to the principles of the Declaration of Helsinki and in accordance with Good Clinical Practice (GCP). The study is registered with ISRCTN registry (ISRCTN48625883).

**Figure 1:**
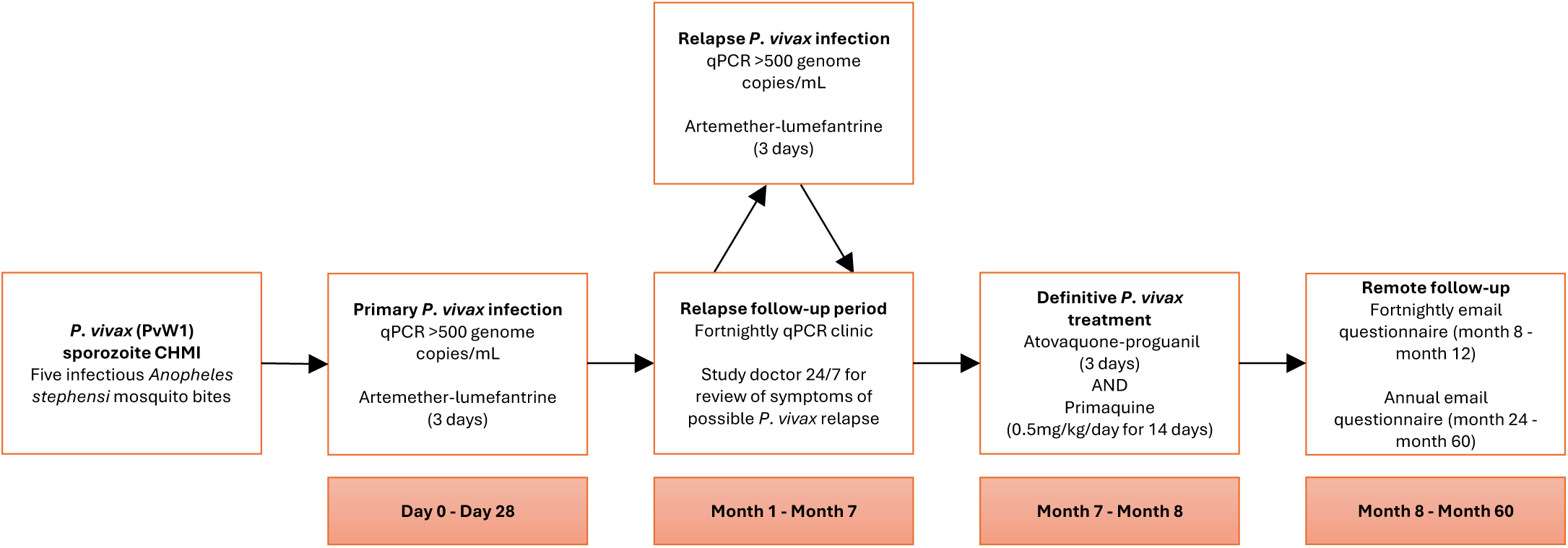
Study design. CHMI = Controlled Human Malaria Infection; qPCR = quantitative polymerase chain reaction

### Procedures

Sporozoite CHMI was administered by five infectious bites of *Anopheles stephensi* mosquitoes infected with PvW1, a clonal isolate of *P. vivax* originating from Thailand (appendix pp 10, 16).^22^ The mosquitoes were infected in a transmission CHMI study (RaViCHMI1, NL-OMON57011) conducted at Radboudumc (Geraedts T *et al.*). In brief, RaViCHMI1 involved a blood-stage CHMI using the cryopreserved PvW1 inoculum administered to healthy adult participants by intravenous injection. Pre-treatment with low-dose mepacrine was used to reduce asexual parasitaemia, while maintaining viable gametocytes, and mosquito feeding took place prior to curative anti-malarial treatment (appendix p 10). The clonal PvW1 stabilate was originally produced in Oxford from a Blood Group O Rh- (O-) donor infected with *P. vivax* by mosquito bite (mosquitoes infected by direct membrane feeding using blood from a source patient in Thailand).^22^

Following sporozoite CHMI, participants received a daily telephone review for 6 days, followed by daily in-person visits from Day 7 for clinical assessment and monitoring of parasitaemia as assessed by quantitative polymerase chain reaction (qPCR) from venous blood sampling. Primary *P. vivax* infection was treated with artemether-lumefantrine (80mg/480mg at 8, 24, 36, 48 and 60 hours) once qPCR > 500 genome copies/mL.

From Day 28 following CHMI, participants entered a six-month follow-up period to monitor for *P. vivax* relapse infections. Participants attended a fortnightly clinic for clinical review and qPCR assessment. Participants were encouraged to contact the study doctor at any time for assessment of possible malaria symptoms arising between clinic visits. If a participant had symptoms suggestive of a *P. vivax* relapse infection, a Relapse Assessment visit (RAx) was performed which included clinical review and qPCR assessment (appendix pp 11–12).

Confirmed *P. vivax* relapse infections (qPCR > 500 genome copies/mL) were treated with artemether-lumefantrine (80mg/480mg at 8, 24, 36, 48 and 60 hours). Treatment was initiated at a Relapse Treatment visit (RTx) which could occur on the same day or the day following the diagnosis of a relapse infection. Participants were reviewed on Day 1, Day 3 and Day 7 following initiation of treatment of primary (T+1, T+3, T+7) and relapse infections (RTx+1, RTx+3, RTx+7).

Definitive anti-malarial treatment was administered at the end of the relapse follow-up period. This comprised of atovaquone-proguanil (1000mg/400mg once daily for three days) followed immediately by primaquine (0·5 mg/kg/day for 14 days). The daily dose of primaquine was rounded up to the nearest whole 15 mg tablet to achieve a total dose >7 mg/kg.^9^ Primaquine treatment was semi-observed, with direct observation or confirmatory telephone call performed on alternate days. Atovaquone-proguanil was used for definitive treatment rather than artemether-lumefantrine due to the inhibition of CYP2D6 by lumefantrine *in vitro* as reported in the Summary of Product Characteristics.^25^ Participants were required to remain local to Oxfordshire from CHMI until completion of primaquine treatment.

Participants attended a final in-person visit 30 days following the start of definitive treatment before undergoing long-term remote follow-up. Email questionnaires were sent fortnightly for four months and will be sent annually until five years (ongoing). These solicit any possible malaria symptoms, and any medical attention sought or received. Participants are encouraged to contact and can be assessed by the study team at any time during this period.

Solicited adverse events (AEs) related to mosquito bites were collected during primary *P. vivax* infection. Solicited AEs related to *P. vivax* infection (foreseeable symptoms of malaria) or anti-malarial or supportive medication were collected during the primary *P. vivax* infection, relapse follow-up period and definitive treatment (appendix p 6). Participants were provided with an oral thermometer.

Unsolicited AEs were collected during the primary *P. vivax* infection, assessment and treatment of relapse infections, and definitive treatment. Medically-attended adverse events and serious adverse events (SAEs) were collected throughout the study. All unsolicited AEs underwent an assessment of severity and causality (appendix p 7–8) and were classified according to the Medical Dictionary for Regulatory Activities (MedDRA version 28.1). An independent Data and Safety Monitoring Committee (DSMC) provided recommendations in the event of any safety concerns raised by the study team.

### Outcomes

The primary objective was to assess the safety, feasibility and frequency of relapsing *P. vivax* infection after experimental CHMI with PvW1 administered by mosquito bite. The co-primary outcomes included safety, as measured by AE occurrences; primary and relapse *P. vivax* infections, as measured by detectable parasitaemia by qPCR +/− clinical symptoms; and the frequency of and time to *P. vivax* relapse infections within a six-month follow-up period. Exploratory outcomes included the serological response to primary and relapsing *P. vivax* PvW1 infection, as measured by enzyme-linked immunosorbent assay (ELISA) to *P. vivax* merozoite surface protein 1 19kDa C-terminal region (PvMSP1-19). Further detailed immunological analyses are ongoing.

### Statistical analysis

Formal sample-size calculations were not performed for this study. The sample size reflects the estimated number of participants required to achieve the primary objective, considering the probability of participants developing primary *P. vivax* infection after CHMI, an estimate of 2–3 relapse infections occurring in a six-month period per participant,^6,11^ and the experience of participant recruitment and retention in previous CHMI studies at the study centre. Data were analysed using R version 4.4.2 and GraphPad Prism 10.6.1.

## Results

Participants were screened for eligibility from 21 January 2025. 20 participants attended in-person screening (figure 2); 11 were provisionally eligible pending CYP2D6 testing. Of these, seven had a predicted “extensive metaboliser” CYP2D6 phenotype, required per protocol to maximise the therapeutic response to primaquine (appendix pp 6, 15). Five participants were enrolled and underwent sporozoite CHMI administered by mosquito bite (five infected mosquitoes per participant) on 15 April 2025. Two participants discontinued the relapse follow-up period early and received definitive anti-malarial treatment at 72 and 154 days following CHMI (one participant required to visit family abroad, one relocated to another area of the UK for study). The remaining three participants received definitive anti-malarial treatment 206 days following CHMI.

**Figure 2:**
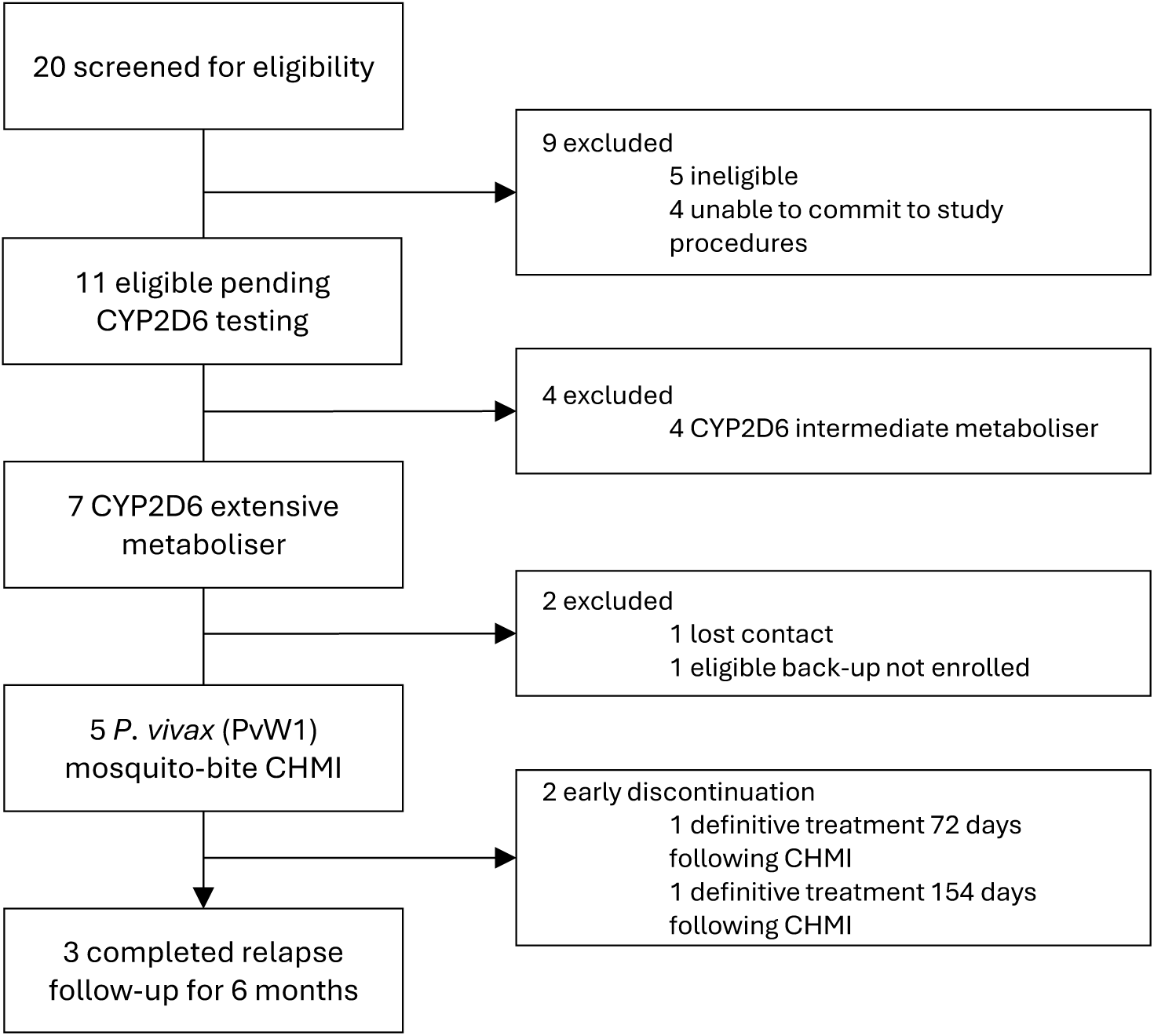
Study profile. CYP2D6 = Cytochrome P450 2D6; CHMI = Controlled Human Malaria Infection; qPCR = quantitative polymerase chain reaction. Participants who completed 6 months of relapse follow-up commenced definitive treatment 206 days following CHMI. Definitive treatment refers to atovaquone-proguanil (3 days) followed by primaquine 0·5 mg/kg/day (14 days).

Baseline characteristics are outlined in table 1. The median age of participants was 22 years (range 20–43 years). Participants included three males (one of whom was assigned female at birth) and two females.

**Table 1:** Baseline characteristics. Data are n or median (range). BMI = body-mass index; CMV = cytomegalovirus; EBV = Epstein-Barr virus.

|  | BIO-006 participants<br>(n=5) |
| --- | --- |
| Sex at birth |  |
| Male | 2 |
| Female | 3 |
| Age (years) | 22<br>(20–43) |
| Weight (kg) | 70·8<br>(52·6–81·6) |
| BMI (kg/m<sup>2</sup>) | 22·9<br>(18·2–27·5) |
| Ethnic origin |  |
| White | 3 |
| Persian | 1 |
| European Asian | 1 |
| CMV seropositive | 3 |
| EBV seropositive | 3 |
| Duffy blood group sero-phenotype |  |
| Fya+b+ | 4 |
| Fya+b- | 1 |
| Fya-b+ | 0 |
| Fya-b- | 0 |

All five participants were diagnosed with a primary *P. vivax* infection 8–10 days following CHMI (figure 3A, appendix p 16) and were treated with artemether-lumefantrine. Parasitaemia was undetectable by qPCR at Day 1, 3 and 7 following initiation of treatment (T+1, T+3, T+7), and at Day 28 following CHMI (C+28) in all participants.

**Figure 3:**
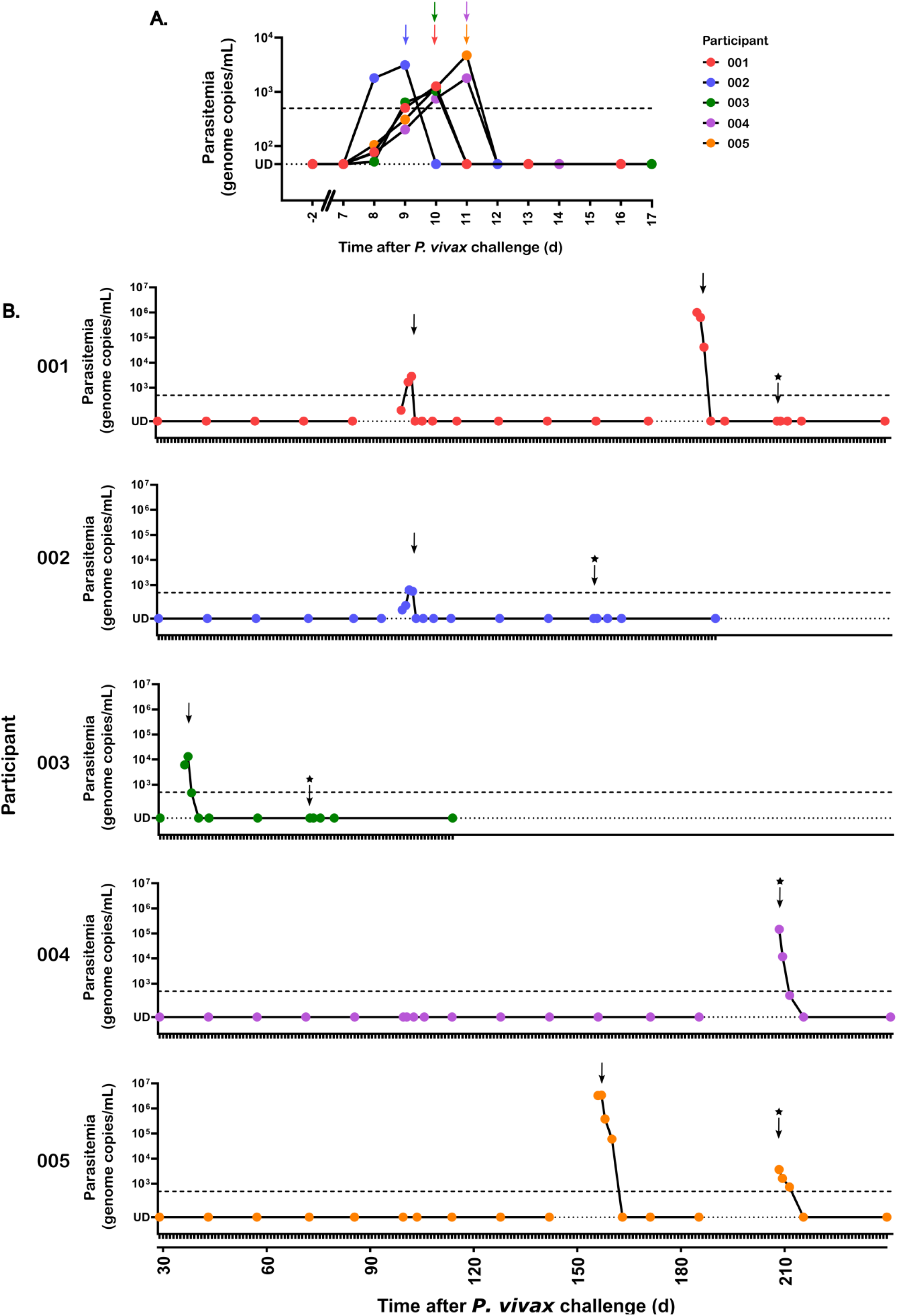
*P. vivax* parasitaemia during primary infection and relapse follow-up period as measured by qPCR. *P. vivax* parasitaemia measured by qPCR in genome copies/mL over time for each participant (n=5) during A) Primary infection and B) Relapse follow-up period. CHMI initiated by mosquito bite on Day 0. Mean of triplicate values within an assay plotted. UD: Undetectable. Dotted line: Lower limit of detection (47 genome copies/mL). Dashed line: Threshold for initiation of treatment (qPCR > 500 genome copies/mL). Arrow: Day of initiation of artemether-lumefantrine (3 days). Arrow with star: Day of initiation of definitive treatment with atovaquone-proguanil (3 days) followed by primaquine 0·5 mg/kg/day (14 days).

Participants subsequently entered the relapse follow-up period. All five participants experienced at least one relapse *P. vivax* infection (qPCR > 500 genome copies/mL); two participants experienced a second relapse infection (figure 3B). First relapse infections were diagnosed at 36, 101, 101, 155 and 206 days following CHMI. The participants who experienced two relapse infections were diagnosed at 101 and 183 days, and 155 and 206 days following CHMI, respectively. Overall incidence rate (from C+28 until definitive treatment) was 3·6 relapse infections per person-year follow-up. Five relapse infections were treated with artemether-lumefantrine (4/5 first, 1/2 second relapses). Two relapse infections were treated with atovaquone-proguanil (1/5 first, 1/2 second relapses) as these were detected on qPCR blood sampling at the initiation of definitive treatment. Parasitaemia was undetectable by qPCR at Day 7 following initiation of treatment of relapse infections in all participants.

Solicited AEs related to mosquito bites were reported in all participants (5/5 redness, 5/5 itching, 3/5 swelling) and were mild (appendix p 19). Solicited AEs related to *P. vivax* infection were mild or moderate during primary and relapse infections (table 2, appendix pp 17–18). Fever > 37·5°C was recorded by 2/5, 2/5 and 1/2 participants at primary infection, first relapse and second relapse, respectively (appendix p 18).

**Table 2:** Solicited adverse events during primary and relapse *P. vivax* infections. Data are n (%) of participants reporting solicited adverse events at primary, first relapse and second relapse *P. vivax* infections. Data show the maximum severity reported by each participant for each adverse event from day qPCR > 500 genome copies/mL (primary infection), or from first presentation with symptoms or qPCR > 500 genome copies/mL (relapse infections) until 7 days following initiation of treatment.

|  | Primary infection<br>(n=5) | First relapse infection<br>(n=5) | Second relapse infection<br>(n=2) |
| --- | --- | --- | --- |
| **Feverishness** |  |  |  |
| Mild | 1 (20%) | 1 (20%) | 0 |
| Moderate | 4 (80%) | 2 (40%) | 1 (50%) |
| Severe | 0 | 0 | 0 |
| **Chills** |  |  |  |
| Mild | 1 (20%) | 1 (20%) | 2 (100%) |
| Moderate | 4 (80%) | 2 (40%) | 0 |
| Severe | 0 | 0 | 0 |
| **Rigor** |  |  |  |
| Mild | 0 | 1 (20%) | 1 (50%) |
| Moderate | 1 (20%) | 0 | 0 |
| Severe | 0 | 0 | 0 |
| **Sweats** |  |  |  |
| Mild | 3 (60%) | 2 (40%) | 0 |
| Moderate | 2 (40%) | 1 (20%) | 0 |
| Severe | 0 | 0 | 0 |
| **Headache** |  |  |  |
| Mild | 3 (60%) | 3 (60%) | 1 (50%) |
| Moderate | 2 (40%) | 1 (20%) | 1 (50%) |
| Severe | 0 | 0 | 0 |
| **Anorexia** |  |  |  |
| Mild | 2 (40%) | 0 | 0 |
| Moderate | 1 (20%) | 1 (20%) | 1 (50%) |
| Severe | 0 | 0 | 0 |
| **Nausea** |  |  |  |
| Mild | 3 (60%) | 2 (40%) | 0 |
| Moderate | 0 | 0 | 1 (50%) |
| Severe | 0 | 0 | 0 |
| **Vomiting** |  |  |  |
| Mild | 2 (40%) | 1 (20%) | 0 |
| Moderate | 0 | 0 | 1 (50%) |
| Severe | 0 | 0 | 0 |
| **Diarrhoea** |  |  |  |
| Mild | 1 (20%) | 0 | 0 |
| Moderate | 0 | 0 | 0 |
| Severe | 0 | 0 | 0 |
| **Myalgia** |  |  |  |
| Mild | 3 (60%) | 2 (40%) | 0 |
| Moderate | 2 (40%) | 0 | 1 (50%) |
| Severe | 0 | 0 | 0 |
| **Arthralgia** |  |  |  |
| Mild | 1 (20%) | 1 (20%) | 0 |
| Moderate | 2 (40%) | 1 (20%) | 1 (50%) |
| Severe | 0 | 0 | 0 |
| **Low back pain** |  |  |  |
| Mild | 2 (40%) | 2 (40%) | 0 |
| Moderate | 1 (20%) | 0 | 2 (100%) |
| Severe | 0 | 0 | 0 |
| **Fatigue** |  |  |  |
| Mild | 1 (20%) | 2 (40%) | 0 |
| Moderate | 3 (60%) | 0 | 2 (100%) |
| Severe | 0 | 0 | 0 |

There were no SAEs during the study. Unsolicited AEs deemed at least possibly related to *P. vivax* infection included one participant with a mild cold sore which started one day prior to diagnosis of first relapse infection, and another participant who experienced severe abdominal pain and severe dehydration in the context of vomiting during treatment of second relapse infection (appendix p 19). This participant was diagnosed with *P. vivax* infection in the absence of symptoms at the definitive treatment visit and received atovaquone-proguanil. Vomiting occurred shortly after administration of the second dose of atovaquone-proguanil (necessitating repeat dosing) and after the third dose. The severe abdominal pain and dehydration started after the third dose and were assessed as possibly due to *P. vivax* infection and probably due to atovaquone-proguanil. The participant was treated with intravenous fluid (Hartmann’s solution 1000mL) and analgesia as an outpatient. Their symptoms significantly improved within a few hours and fully resolved in the following days. The participant remained afebrile throughout this episode. In consultation with the Chair of the DSMC, it was agreed to postpone primaquine treatment for this participant by one week, after which a full treatment course was completed without incident.

One participant presented with isolated cyanosis of the lips on Day 11 following the initiation of definitive treatment (Day 9 of primaquine treatment). They were assessed and diagnosed with primaquine-induced methaemoglobinaemia with a methaemoglobin (MetHb) level of 11%. In consultation with the DSMC and a local haematology specialist at Oxford University Hospital NHS Foundation Trust, it was agreed to discontinue primaquine treatment and monitor the participant as an outpatient. The participant remained stable during this period (SaO2 ≥ 94%) with no symptoms of shortness of breath and MetHb decreased to 7·6% within four days. By Day 30 following the initiation of definitive treatment, MetHb was normal (1·5%). In consultation with the DSMC, it was agreed not to resume primaquine treatment as it was expected that the participant had received a fully effective therapeutic dose (total dose 5·3 mg/kg). The total dose of primaquine received prior to discontinuation was comparable to primaquine 30mg once daily for 14 days (total dose 5·5 mg/kg) which is recommended in many non-weight-based guidelines.^26^ A genetic screen for congenital causes of methaemoglobinaemia was performed; no pathogenic variants were detected.

Other medication-related AEs included mild palpitations (artemether-lumefantrine, primary infection), moderate diarrhoea (atovaquone-proguanil, definitive treatment) and mild nausea (primaquine, definitive treatment) which occurred in different participants (appendix p 20). The participant with palpitations had accidentally taken an extra third dose of artemether-lumefantrine within 12 hours on the first day of treatment of primary infection. Their electrolytes were checked on Day 1 following initiation of treatment and were normal. They subsequently completed the full treatment course in addition to the extra dose. The same participant developed Grade 3 hypokalaemia (potassium 3·0 mmol/L) at Day 3 following initiation of treatment which resolved within 48 hours. This was considered to be possibly related to the study medication, but more likely related to *P. vivax* infection due to mild diarrhoea. The palpitations occurred in the context of significant caffeine intake and a prior history of palpitations, and were considered unlikely to be related to hypokalaemia due to relatively mild change from baseline. An electrocardiogram was normal (QTc 443).

Laboratory AEs at primary and relapse *P. vivax* infections included anaemia, leukopenia, neutropenia, lymphopenia, thrombocytopenia, hypokalaemia, elevated urea and elevated ALT (appendix pp 21–22). These included the following five Grade 3 abnormalities: One participant experienced Grade 3 thrombocytopenia at primary and first relapse infection. This participant had mild thrombocytopenia (Grade 1) at baseline, and, in each instance, platelets returned to baseline within six days (appendix p 22). One participant experienced Grade 3 decrease in haemoglobin at first relapse infection. This was a decrease in haemoglobin to 133 g/L (Day 3 following initiation of atovaquone-proguanil treatment) from a baseline of 157 g/L. Haemoglobin returned to 144 g/L within four days. One participant experienced Grade 3 lymphopenia at second relapse infection. This occurred concurrently with maximum severity of symptoms of severe abdominal pain, severe dehydration and vomiting (see above) at Day 2 following initiation of atovaquone-proguanil treatment, and returned to normal within 24 hours (appendix p 22). One participant experienced Grade 3 hypokalaemia as described previously.

The circumstances surrounding the presentation of relapse infections are summarised in the appendix (p 28). 3/5 participants experienced an upper respiratory tract infection in the week prior to presentation with a relapse infection. 4/5 participants experienced higher parasitaemia at first relapse infection compared to primary infection (appendix p 23); although the primary infection was monitored by daily qPCR, likely resulting in earlier detection and treatment compared to relapse infections. Maximum detected parasitaemia by qPCR was greater at relapse infections (median 13,289, range 639–3,357,823 genome copies/mL) compared to primary infection (median 1,796, range 1,042–4,719 genome copies/mL) although this was not statistically significant (Wilcoxon signed-rank test, p = 0·15).

Anti-PvMSP1-19 total IgG was detectable in 1/5 participant at Day 7 following treatment of primary infection. All participants (5/5) seroconverted following first relapse infection with detectable anti-PvMSP1-19 IgG responses which were maintained until 30 days following initiation of definitive anti-malarial treatment (figure 4).

**Figure 4:**
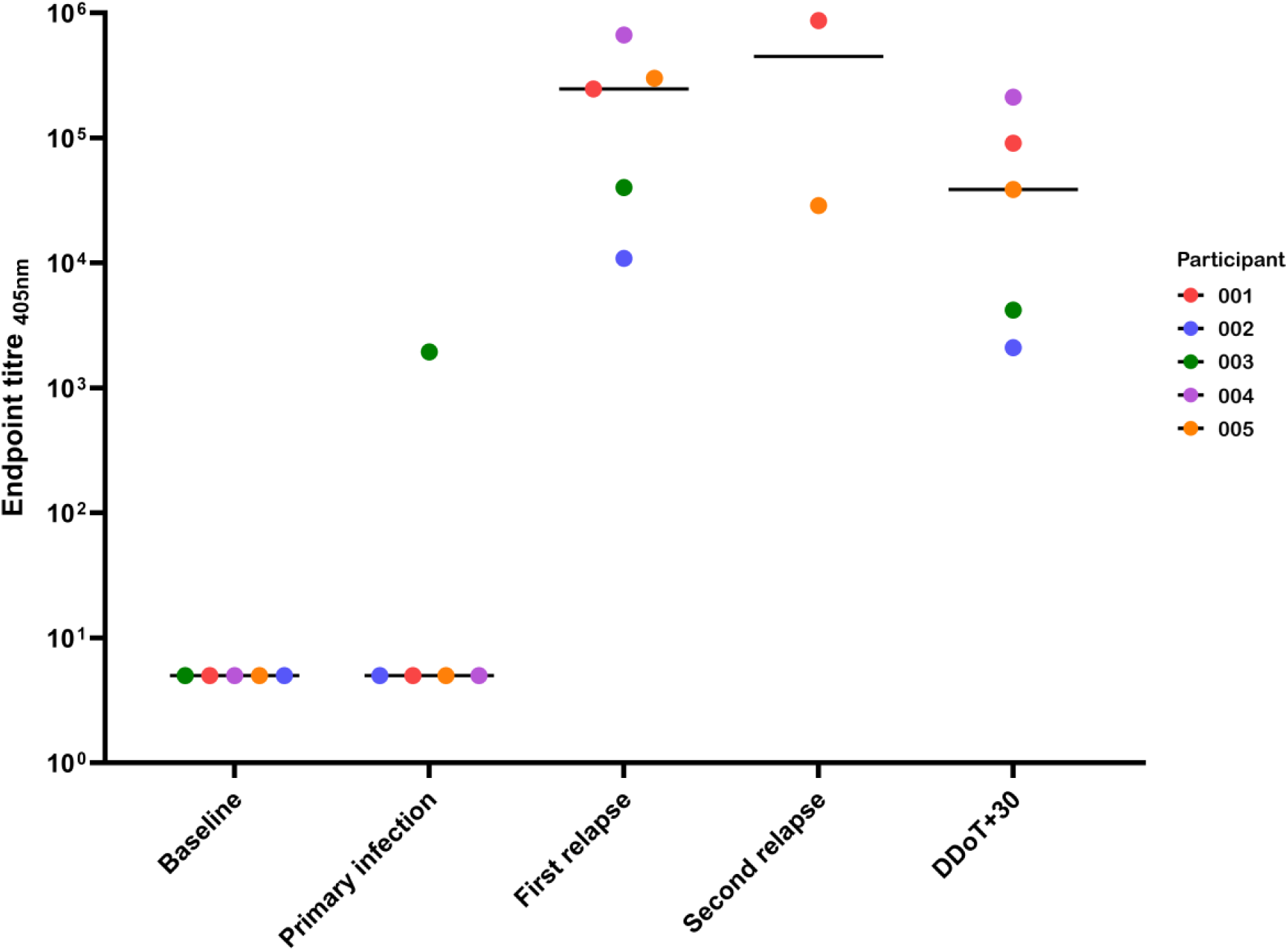
Antibody responses to Merozoite Surface Protein 1 (MSP1-19) during primary infection and relapse follow-up period. Median and individual anti-PvMSP1-19 total IgG responses as measured by endpoint ELISA for each participant. The following timepoints are shown: Baseline = 2 days prior to CHMI (n=5); Primary infection = 7 days following treatment of primary infection (n=5); First relapse = 7 days following treatment of first relapse infection (n=5); Second relapse = 7 days following treatment of second relapse infection (n=2); DDoT+30 = 30 days following day of initiation of definitive anti-malarial treatment (n=5). Bar indicates median.

There was no clear association with local climate conditions and relapse infections (appendix p 29). Mean daily temperature from CHMI to definitive treatment was 15·8°C and mean weekly rainfall was 9·4mm.

Long-term remote follow-up is ongoing. To date, more than six months after the initiation of definitive treatment, no further *P. vivax* relapse infections have been recorded.

## Discussion

Here, we report the safety and feasibility of a CHMI model for relapsing *P. vivax* using the clonal isolate PvW1 administered by mosquito bite. Five participants with normal blood G6PD levels and extensive CYP2D6 metaboliser phenotype underwent CHMI. All participants developed primary *P. vivax* infection and experienced at least one relapse infection in a six-month follow-up period; attack rate 100% (5/5). Two participants experienced a second relapse prior to the administration of primaquine treatment. Solicited AEs were mild or moderate and there were no SAEs.

Experimental infection of humans with *P. vivax* has been performed for over 125 years.^6^ The earliest studies provided evidence of the mosquito-malaria transmission theory. A remarkable period of medical history followed in the early 20^th^ Century in the form of malariotherapy for the treatment of neurosyphilis.^6^ Thousands of patients received *Plasmodium* infection as a treatment, and strains of *P. vivax* (e.g. Madagascar strain) were propagated from patient to patient. The practice of malariotherapy was ultimately discontinued following the advent of antibiotic therapy. Nevertheless, this period continues to be informative for modern CHMI studies.

In the modern era, *P. vivax* mosquito-bite CHMI studies have been conducted using mosquitoes infected from patients with naturally acquired infections.^18^ This creates challenges for reproducibility and interstudy comparison as the parasite strain inevitably differs between each study. Although cross-strain protection is a desirable quality of malaria vaccines, harnessing a reproducible mosquito-bite CHMI model with a single clonal isolate would facilitate early down-selection of pre-erythrocytic vaccine candidates. However, this has been challenging to achieve due to an inability to sustain long-term *in vitro* cultures of *P. vivax* parasites and thus gametocyte production needed for the generation of laboratory-infected mosquitoes. In our study, we used mosquitoes infected with PvW1 by feeding on blood from participants in a preceding blood-stage CHMI study (Geraedts T *et al.*). In doing so, we have successfully completed the full lifecycle of PvW1 and demonstrated a mosquito-bite CHMI using a well-characterised clonal isolate.

*P. vivax* relapse infections have been reported in previous CHMI studies. In 1901, Dr Patrick Thurburn Manson experienced a relapse infection nine months after participating in an experimental *P. vivax* infection study conducted by his father, Sir Patrick Manson.^27^ More recently, two participants experienced relapses after primaquine treatment during a Phase 1/2a VMP001/AS01_B_ vaccine CHMI study at the Walter Reed Army Institute of Research.^11^ The mosquitoes in this study were infected from a source patient in Thailand. One participant experienced two relapse infections (9 and 18 weeks following CHMI) and the other experienced three relapses (11, 20 and 48 weeks following CHMI). Retrospective CYP2D6 testing demonstrated that these participants had intermediate (CYP2D6 *4 / *41) and poor metaboliser (CYP2D6 *5 / *6ABC) phenotypes respectively, providing one of the earliest indications of primaquine failure due to CYP2D6 polymorphisms.^11^ Participants in this study were initially treated with primaquine 30 mg once daily for 14 days. Relapses were treated with a higher primaquine dose based on weight (total dose > 6 mg/kg). These participants would not have been eligible for inclusion in our current study due to extensive CYP2D6 testing performed at screening. Additional factors to optimise the effectiveness of primaquine in our study included high-dose treatment (total dose > 7 mg/kg)^9^ and observation of therapy. Long-term follow-up of participants is ongoing. However, to date, more than six months after primaquine treatment, no relapse infections have been recorded.

The mechanisms responsible for hypnozoite reactivation are unknown.^6,7^ One proposal is that the parasite may have an “internal clock”, evolved through natural selection to suit local seasonal conditions. In tropical regions, relapses appear to occur frequently (∼30–45 days) while in temperate climates there may be a longer latent period.^3,6^ In our study, using PvW1 originating from Thailand, relapse frequency was variable between participants. The earliest relapse was detected at 36 days and the latest first relapse was detected at 206 days following CHMI. On two occasions, we diagnosed relapse infections in two participants on the same day (101 and 206 days following CHMI). Although influenced by our fortnightly sampling frequency, relapses in these individuals nevertheless occurred within the same 2-week period. Blood-stage anti-malarial treatment is recognised to provide a degree of post-treatment prophylaxis and may suppress early relapse infections.^6^ The elimination half-life of lumefantrine is 2–6 days. Therefore, participants in our study likely had a degree of therapeutic drug cover in the weeks following artemether-lumefantrine treatment which may confound true relapse frequency.^25^ However, one participant had a relapse detected 36 days following CHMI (23 days after completing artemether-lumefantrine treatment).

An alternative (or complementary) theory of hypnozoite activation is that it may be triggered by external factors.^6,7^ These may “indicate” conditions are favourable for mosquito transmission and may include environmental changes (e.g. temperature, rainfall, day length), mosquito bites themselves, or other febrile illnesses. A disproportionate number of *P. vivax* relapse infections occur following *P. falciparum* infection.^6,7,28^ In our study, 3/5 participants experienced symptoms of an upper respiratory tract infection in the week prior to relapse infection, one of these participants also had a cold sore. However, our sample size is too small to comment definitively on potential triggers. There were also reports of viral upper respiratory tract symptoms by participants (including one confirmed diagnosis of COVID-19) who did not experience a relapse infection in the following days or weeks.

Regular qPCR testing and a low threshold for treatment (qPCR > 500 genome copies/mL) meant that anti-malarial medication was initiated early during *P. vivax* infections, often prior to onset of symptoms. In addition, the different frequency of qPCR monitoring between primary (daily qPCR) and relapse infections (fortnightly qPCR and if symptomatic) confounds comparisons between primary and relapse infections. The maximum recorded parasitaemia was typically higher at relapse infections, likely due to less frequent sampling and thus later detection. Nevertheless, symptoms of malaria (solicited AEs) at primary and relapse infections were similar; here a degree of acquired immunity may have played a role. In a previous repeat blood-stage CHMI study using PvW1 (qPCR treatment threshold 10,000 genome copies/mL), we demonstrated that clinical immunity to *P. vivax* is acquired rapidly after a single infection.^24^ However, in this previous study, all participants seroconverted to PvMSP1-19 after primary infection while, in the current study, only 1/5 participants seroconverted to PvMSP1-19 following primary infection, likely due to the lower treatment threshold and the shorter exposure of the immune system to blood-stage parasites. All participants seroconverted to PvMSP1-19 following first relapse infection.

Although solicited AEs associated with *P. vivax* infections were typically mild or moderate, our study raised two important safety considerations. One participant experienced severe abdominal pain and dehydration in the context of vomiting requiring treatment with intravenous fluids. These symptoms occurred two days following the diagnosis of their second relapse infection and were deemed most likely due to atovaquone-proguanil. Atovaquone-proguanil was used for definitive treatment (rather than artemether-lumefantrine) as lumefantrine inhibits CYP2D6 *in vitro*.^25^ However, atovaquone-proguanil is associated with gastrointestinal side effects. Another participant experienced primaquine-induced methaemoglobinaemia, presenting with cyanosis of the lips on Day 9 of primaquine treatment. Their primaquine treatment regimen was curtailed (total dose 5·3 mg/kg). Methaemoglobinaeamia is proposed as a surrogate marker of treatment efficacy as, for a given primaquine regimen, higher methaemoglobin levels correlates with a reduced risk of future relapse.^10^ These instances are informative for future relapse CHMI studies, and ultimately highlight the imperfections of current anti-malarial therapies.

This study has limitations. First, we only recruited five healthy UK adults (median age 22 years) with normal G6PD levels and extensive CYP2D6 metaboliser phenotype with no history of prior malaria infection; utilising this model in endemic populations would be of significant value in the future. Second, although CHMI administered by mosquito bite is the method most representative of naturally-acquired infection, we used a five bite protocol with *A. stephensi* mosquitoes to maximise the likelihood of successful infection. This potentially results in a high sporozoite load which may influence hypnozoite burden and relapse frequency;^6^ varying the number of mosquito bites to explore how this impacts relapse frequency would be of substantial interest. Finally, participants were also monitored for relapse infections in a geographical setting where malaria transmission does not occur; this minimises risk of onward transmission but environmental triggers are likely to be different in endemic areas.

To our knowledge, we report the first CHMI study in the modern era to administer a well-characterised clonal isolate of *P. vivax* (PvW1) by mosquito bite. We have also demonstrated a CHMI model for relapsing *P. vivax* with a reproducible algorithm (appendix pp 11–12) for the safe monitoring of participants over a six-month relapse follow-up period. There are no licensed vaccines for *P. vivax* malaria. An effective pre-erythrocytic *P. vivax* vaccine could significantly reduce the incidence of relapse infections, potentially enough to interrupt transmission in certain settings.^29^ Hypnozoiticidal drugs are crucial in countries striving for *P. vivax* elimination. However, novel therapies are required as the current 8-aminoquinolines are imperfect due to contraindications, side effects and variable efficacy. Although careful consideration is required to minimise and manage side effects of anti-malarial medications, and protect study participants from primaquine treatment failure, we have demonstrated that a CHMI model for relapsing *P. vivax* is safe and feasible. This proof-of-concept study presents an opportunity to explore the mechanisms behind hypnozoite formation and activation, and provides a platform for future drug and vaccine efficacy studies against *P. vivax*.

## Data Availability

Data associated with this study are present in the paper or appendix and will be available after the end of the study upon reasonable request that should be directed to the corresponding author. Proposals will be reviewed and approved by the Sponsor, Chief Investigator, and collaborators. After approval of a proposal, data can be shared through a secure online platform after signing a data access agreement. Any shared data will be de-identified.

## Contributors

ADSD, BM, FRD, SES, SJD and AMM conceived the trial. AMM was the chief investigator and SJD was the senior laboratory investigator. ADSD, TJMG, DM, ECB, JKvH, TWR, CAR, JS, MMH, CM, JB, CN, EC, YFM, BGW, KM, ZWT, ASR, CAG, HR, EP, MBBM, TB, MG, JB, WG, G-JvG, CMN, BM, FRD, SES, SJD and AMM contributed to the implementation of the study and data collection. ADSD, ECB, FRD, SES, SJD and AMM analysed the data. KK, REC, AB, PE, NO, J-SC, FLN performed project management. ADSD, FRD, SES, SJD and AMM interpreted the data and contributed to writing the manuscript. All authors had full access to all the data in the study and had final responsibility for the decision to submit for publication.

## Acknowledgements

We thank all the BIO-006 study participants (including those not enrolled) without whom this research would not be possible. We also thank the independent DSMC, Ruth Payne, Quirijn de Mast and Johannes Mischlinger, for overseeing the trial; Noémi Roy for providing real-time advice on management of methaemoglobinaemia; Charlie Firth, Isil Senol and Laura Borg for communications and public engagement; Andrew Roberts for Quality Assurance; Stella Mwakio and Azeezat Ajose for monitoring; the Oxford Vaccine Group clinical team for support with study visits and on-call cover; all the partners of the OptiViVax consortium (including independent scientific advisory committee) for their support and input. OptiViVax is co-funded from the European Union Horizon Europe programme under grant agreement No. 101080744. The project also receives funding from UK Research and Innovation (UKRI) under the UK government’s Horizon Europe funding guarantee (grant No. 10077974) and Swiss Government’s State Secretariat for Education, Research, and Innovation (SERI No. 23.00182). Views and opinions expressed are those of the author(s) only and do not necessarily reflect those of the European Union, UK Research and Innovation or Swiss Government’s State Secretariat for Education, Research, and Innovation. Neither the European Union, UK Research and Innovation or Swiss Government’s State Secretariat for Education, Research, and Innovation nor the granting authority can be held responsible for them. The study was also funded in part by the UK National Institute for Health and Care Research (NIHR) Biomedical Research Centre: Oxford. The views expressed are those of the authors and not necessarily those of the National Health Service, the NIHR, or the Department of Health. JS holds a UK Medical Research Council Clinical Research Training Fellowship [MR/Z505134/1].

