## Supplementary Appendix for "A Controlled Human Malaria Infection model for relapsing *Plasmodium vivax*"

### Supplementary Methods

#### Inclusion criteria for the BIO-006 study

The participant must satisfy all the following criteria to be eligible for the study:

- Healthy, malaria-naïve adult aged 18 to 45 years.
- Able and willing to provide informed consent to participate in the study.
- Able and willing (in the opinion of the Investigator) to comply with all study requirements.
- Willing to allow the Investigators to access participant's electronic medical records or discuss the participant's s medical history with their GP.
- Participants of childbearing potential only: must practice continuous highly effective contraception until 3 months after completion of Primaquine treatment
- Negative haemoglobinopathy screen (including sickle cell disease and alpha and beta thalassaemia).
- Normal G6PD screen.
- Agreement to refrain from blood donation until at least 3 years following completion of Primaquine treatment, as per current UK Blood Transfusion and Tissue Transplantation Services guidelines.
- Able to answer all questions on the informed consent questionnaire correctly at first or second attempt.
- Able to travel to CCVTM easily.
- Able to travel to the Netherlands for malaria challenge with the necessary passport +/- visa requirements.
- Reachable 24 hours a day by mobile phone during the period between CHMI and completion of Primaquine treatment.
- Willing to take anti-malarial treatment for i) primary *P. vivax* infection ii) any relapse *P. vivax* infections and iii) at the end of relapse follow-up period as outlined in schedule of study procedures.
- Willing to remain in Oxfordshire (or surrounding area) following malaria challenge (after return from the Netherlands) until completion of treatment of primary *P. vivax* infection.
- Willing to remain within travelling distance of Oxfordshire (or surrounding area) following treatment of primary *P. vivax* infection until completion of Primaquine treatment (month 7). Absolute necessity is to remain on the UK mainland within 1-2 hours of secondary care NHS hospital.
- Willing to be registered on the TOPS database (The Over volunteering Prevention System; www.tops.org.uk).

#### Exclusion criteria for the BIO-006 study

The participant may not enter the study if ANY of the following apply:

- Red blood cells negative for the Duffy antigen/chemokine receptor (DARC).
- CYP2D6 genotype suggestive of poor or intermediate metabolism of Primaquine.
- Body weight <50kg or Body Mass Index (BMI) <18.0 at screening.
- History of clinical malaria (any species) or previous participation in any malaria vaccine trial or CHMI.
- Travel to a clearly malaria endemic locality during the in-person study period or within the preceding six months.

- Receipt of immunoglobulins or blood products (e.g. blood transfusion) in the last three months.
- Receipt of an investigational product in the 30 days preceding enrolment (or planned receipt during the study period) likely to impact on interpretation of the trial data or the *P. vivax* parasite as assessed by the Investigator.
- Concurrent involvement in another clinical trial involving an investigational product or planned involvement during the study period.
- Any confirmed or suspected immunosuppressive or immunodeficient state, including HIV infection; asplenia; recurrent, severe infections and chronic (more than 14 days) immunosuppressant medication within the past 6 months (inhaled and topical steroids are allowed).
- Any history of severe allergy or anaphylaxis.
- Use of systemic antibiotics with known anti-malarial activity within 30 days of CHMI (e.g. trimethoprim-sulfamethoxazole, doxycycline, tetracycline, clindamycin, erythromycin, fluoroquinolones and azithromycin).
- Use of anti-malarials within 30 days of CHMI.
- Any clinical condition known to prolong the QT interval.
- History of cardiac arrhythmia, including clinically relevant bradycardia.
- Disturbances of electrolyte balance, e.g. hypokalaemia or hypomagnesaemia.
- Family history of congenital QT prolongation or sudden death.
- An estimated ten-year risk of fatal cardiovascular disease of ≥5% at screening, as determined by the Systematic Coronary Risk Evaluation (SCORE2).
- Use of medications known to have a potentially clinically significant interaction with both Riamet and Malarone.
- Use of medications known to have a potentially clinically significant interaction with Primaquine.
- Any other contraindications/known hypersensitivities to both Riamet and Malarone.
- Any other contraindications/known hypersensitivities to Primaquine.
- History of sickle cell anaemia, sickle cell trait, thalassaemia or thalassaemia trait, G6PD deficiency or any haematological condition that could affect susceptibility to malaria infection.
- Pregnancy, lactation or intention to become pregnant during the study.
- History of cancer (except basal cell carcinoma of the skin and cervical carcinoma in situ).
- History of serious psychiatric condition that may affect participation in the study.
- Any other serious chronic illness requiring hospital specialist supervision.
- Suspected or known current alcohol misuse.
- Suspected or known injecting drug use in the 5 years preceding enrolment.
- Hepatitis B surface antigen (HBsAg) detected in serum.
- Seropositive for hepatitis C virus (antibodies to HCV) at screening (unless participant has taken part in a prior hepatitis C vaccine study with confirmed negative HCV antibodies prior to participation in that study, and negative HCV ribonucleic acid (RNA) qPCR at screening for this study).
- Participants unable to be closely followed for social, geographic or psychological reasons.

- Any clinically significant abnormal finding on biochemistry or haematology blood tests, or clinical examination.
- Any other significant disease, disorder, or finding (in the opinion of the Investigator) which may significantly increase the risk to the volunteer because of participation in the study, affect the ability of the volunteer to participate in the study or impair interpretation of the study data.
- Inability of the study team to confirm medical history via electronic records or contact the participant's GP to confirm medical history.

#### CYP2D6 testing

CYP2D6 testing was performed on venous blood samples of participants who were provisionally eligible for inclusion (pending CYP2D6 result) following in-person screening. Testing was performed by Immunodiagnostik AG and reported by SDS Special Diagnostic Services GmBH. The following single nucleotide polymorphisms (SNPs) were included: CYP2D6*2 (886C>T), CYP2D6*2 (1457G>C), CYP2D6*3 (2549delA), CYP2D6*4 (1846G>A), CYP2D6*5 (Gene Deletion), CYP2D6*6 (1707delT), CYP2D6*7 (2935A>C), CYP2D6*8 (1758G>T), CYP2D6*9 (2615_2617delAAG), CYP2D6*10 (100C>T), CYP2D6*11 (883G>C), CYP2D6*12 (124G>A), CYP2D6*17 (1023C>T), CYP2D6*29 (3183G>A), CYP2D6*35 (31G>A), CYP2D6*41 (2988G>A), CYP2D6*43 (77G>A), CYP2D6*45 (1717G>A), CYP2D6*46 (77G>A), CYP2D6*46 (1717G>A) and CYP2D6*xN (Gene Duplication). Detected SNPs were reported as homozygous or heterozygous. Results included a predicted CYP2D6 genotype and CYP2D6 phenotype (poor, intermediate, extensive or ultra-rapid metaboliser). CYP2D6 phenotype was according to resources available at ClinPGx (clinpgx.org).[1] Participants were not eligible for inclusion if they had a poor or intermediate CYP2D6 metaboliser phenotype.

#### Solicited adverse events

Solicited adverse events (AEs) refer to the foreseeable symptoms, signs and laboratory findings which may occur following a study intervention. This includes i) following mosquito bites, or ii) following *P. vivax* infection (either primary or relapse), or iii) following anti-malarial or supportive medications provided by the study team.

Solicited AEs related to mosquito bites include itch, redness, swelling and warmth.

Solicited AEs related to malaria infection include feverishness, chills, rigor, sweats, headache, anorexia, nausea, vomiting, diarrhoea, myalgia, arthralgia, low back pain, fatigue, fever, tachycardia, hypotension, lymphopenia and thrombocytopenia.

Solicited AEs related to anti-malarial or supportive medication (paracetamol and cyclizine) provided by the study team are listed as undesirable effects in the Summary of Product Characteristics (SmPCs) for these medications. These were solicited while the participant is taking this medication.

#### Serious adverse events and medically attended adverse events

A serious adverse event is defined as any untoward medical occurrence that:

- results in death
- is life-threatening
- requires inpatient hospitalisation or prolongation of existing hospitalisation
- results in persistent or significant disability/incapacity
- consists of a congenital anomaly or birth defect.

A medically attended adverse event refers to any untoward medical occurrence that led to assessment by a healthcare provider.

#### Causality assessment of adverse events

For every unsolicited AE, an assessment of the relationship of the AE to the study intervention was undertaken. The "study intervention" is considered sporozoite *P. vivax* PvW1 CHMI and, for the purpose of AE causality assessment, includes i) mosquito bites, ii) *P. vivax* infection (either primary or relapse), and iii) anti-malarial or supportive medications provided by the study team. Alternative causes of the AE, such as the natural history of pre-existing medical conditions, concomitant therapy, other risk factors and the temporal relationship of the event to CHMI were considered. The likely causality of unsolicited AEs was assessed as per the criteria below:

No Relationship: No temporal relationship to study intervention ***and*** alternate aetiology (clinical state, environmental or other interventions); ***and*** does not follow known pattern of response to CHMI.

Unlikely: Unlikely temporal relationship to study intervention ***and*** alternate aetiology likely (clinical state, environmental or other interventions) ***and*** does not follow known typical pattern of response to CHMI.

Possible: Reasonable temporal relationship to study intervention; ***or*** event not readily produced by clinical state, environmental or other interventions; ***or*** similar pattern of response to that known to occur following CHMI.

Probable: Reasonable temporal relationship to study intervention; ***and*** event not readily produced by clinical state, environment, or other interventions ***or*** known pattern of response to that known to occur following CHMI.

Definite: Reasonable temporal relationship to study intervention; ***and*** event not readily produced by clinical state, environment, or other interventions; ***and*** known pattern of response to that known to occur following CHMI.

### Severity grading of adverse events

Participant reported AEs were graded as mild, moderate or severe according to the following criteria:

Grade 1: Transient or mild discomfort (< 48 hours); no medical intervention/therapy required.

Grade 2: Mild to moderate limitation in activity, some assistance may be needed; no or minimal medical intervention/therapy required.

Grade 3: Marked limitation in activity, some assistance usually required; may require medical intervention/therapy.

**Table S1: Severity grading for clinically significant physical observations**

| | **Grade 1** | **Grade 2** | **Grade 3** |
|---|---|---|---|
| Tachycardia – beats per min | 101–115 | 116–130 | >130 |
| Hypotension (systolic) mm Hg | 85–89 | 80–84 | <80 |
| Hypertension (systolic) mm Hg | 141–159 | 160–179 | ≥180 |
| Hypertension (diastolic) mm Hg | 91–99 | 100–109 | ≥110 |
| Fever °C | 37·6 – 38·0 | 38·1-39·0 | >39·0 |

**Table S2: Severity grading for clinically significant laboratory abnormalities**

| | **Grade 1** | **Grade 2** | **Grade 3** |
|---|---|---|---|
| Haemoglobin: decrease from baseline value (g/L) | 10 – 15 | 16 – 20 | > 20 |
| White cell count: Elevated (x $10^9$/L) | 11·10 – 15·00 | 15·01 – 20·00 | > 20·00 |
| White cell count: Depressed (x $10^9$/L) | 2·50 – 3·50 | 1·50 – 2·49 | < 1·50 |
| Neutrophil count (x $10^9$/L) | 1·50 – 1·69 | 1·00 – 1·49 | < 1·00 |
| Lymphocyte count (x $10^9$/L) | 0·75 – 0·89 | 0·50 – 0·74 | < 0·50 |
| Eosinophil count (x $10^9$/L) | 0·65 – 1·50 | 1·51 – 5·00 | > 5·00 |
| Platelet count (x $10^9$/L) | 125 – 149 | 100 – 124 | < 100 |
| Sodium: hyponatraemia (mmol/L) | 132 – 134 | 130 – 131 | < 130 |
| Sodium: hypernatraemia (mmol/L) | 146 | 147 | > 147 |
| Potassium: hypokalaemia (mmol/L) | 3·3 – 3·4 | 3·1 – 3·2 | < 3·1 |
| Potassium: hyperkalaemia (mmol/L) | 5·4 – 5·5 | 5·6 – 5·7 | > 5·7 |
| Urea (mmol/L) | 8·2 – 9·3 | 9·4 – 11·0 | > 11·0 |
| Creatinine (µmol/L) | 132 – 150 | 151 - 177 | > 177 |
| ALT (IU/L) | 50 – 112 | 113 – 229 | > 229 |
| AST (IU/L) | 46 – 105 | 106 – 213 | > 213 |
| Bilirubin, with increase in LFTs (µmol/L) | 23 – 25 | 26 – 31 | > 31 |
| Bilirubin, with normal LFTs (µmol/L) | 23 – 33 | 34 – 41 | > 41 |
| ALP (IU/L) | 143 – 272 | 273 – 402 | > 402 |
| Albumin (g/L) | 28 – 31 | 25 – 27 | <25 |

ALT = Alanine transaminase, AST = Aspartate transaminase, ALP = Alkaline phosphatase

#### Infection of mosquitoes (RaViCHMI1 study)

RaViCHMI1 was a CHMI study conducted at Radboud University Medical Center (Radboudumc) with the primary objective of reproducibly infecting mosquitoes with blood from human participants undergoing blood-stage *P. vivax* CHMI using PvW1 administered intravenously (Geraedts T *et al.*). RaViCHMI1 is summarised below as it was the source of the infected mosquitoes used in this study.

RaVICHMI1 participants were recruited sequentially into three cohorts (four participants in each). Mosquito infection rate was maximised using a combination of *ex vivo* gametocyte enrichment techniques and gametocyte-sparing anti-malarial treatment to attenuate malaria symptoms and reduce asexual parasitaemia (without affecting gametocytes) prior to the administration of curative anti-malarial treatment with atovaquone-proguanil. These techniques were optimised over the three cohorts.

*Anopheles stephensi* mosquitoes from RaViCHMI1 Cohort 3 participants were used for sporozoite CHMI in this study. Cohort 3 participants were screened for HIV, Hepatitis B, Hepatitis C, West Nile virus and other mosquito-borne diseases if indicated by travel history prior to enrolment. Asexual PvW1 parasites were administered intravenously. Cohort 3 participants received mepacrine as gametocyte-sparing treatment (100mg every 8 hours on the first day, then 100mg once daily for 3 days) once parasitaemia was greater than 10 parasites/µL on Thick Blood Smear (TBS) microscopy or if they had any density of parasitemia with either fever (>38·5°C) or malaria symptoms interfering with daily activities. Infection of mosquitoes was performed using Direct Skin Feeding (DSF) and Direct Membrane Feeding Assay (DMFA) with and without enrichment of gametocytes (65% Percoll and centrifugation at 1500xg). Curative treatment was administered on Day 22 following CHMI once all mosquito feeding assays had been completed. The blood-stage CHMI for RaViCHMI Cohort 3 participants was performed on 13 March 2025.

### Treatment algorithms for relapse *P. vivax* infections

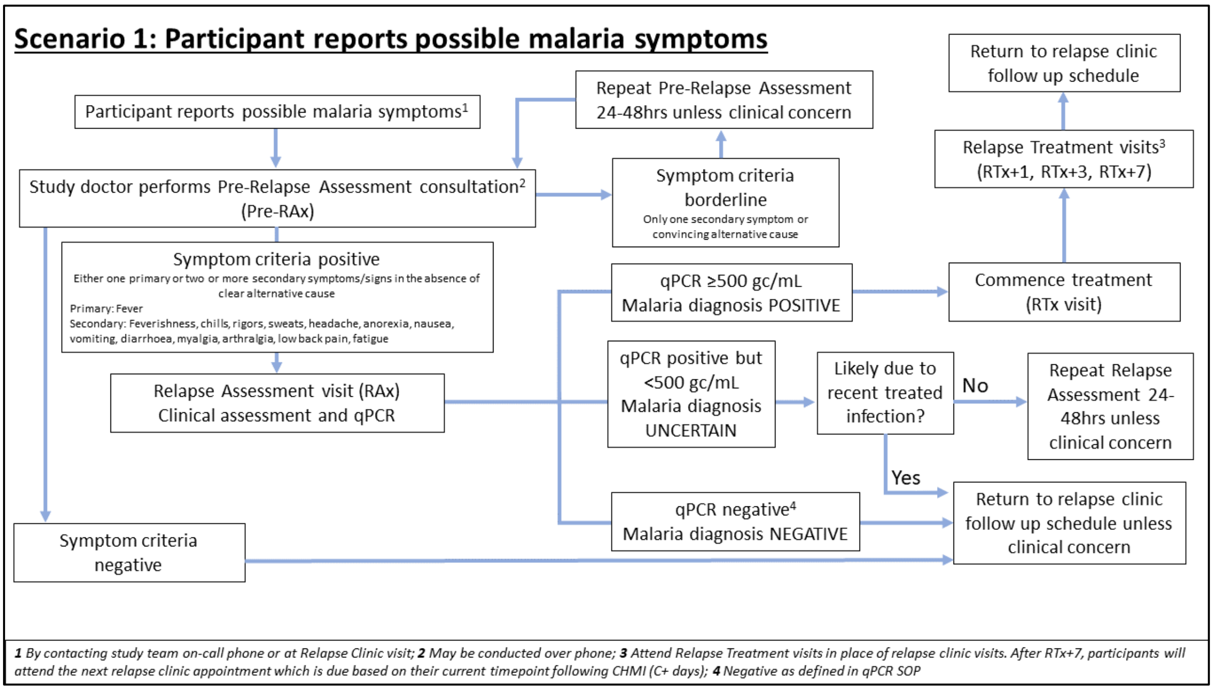

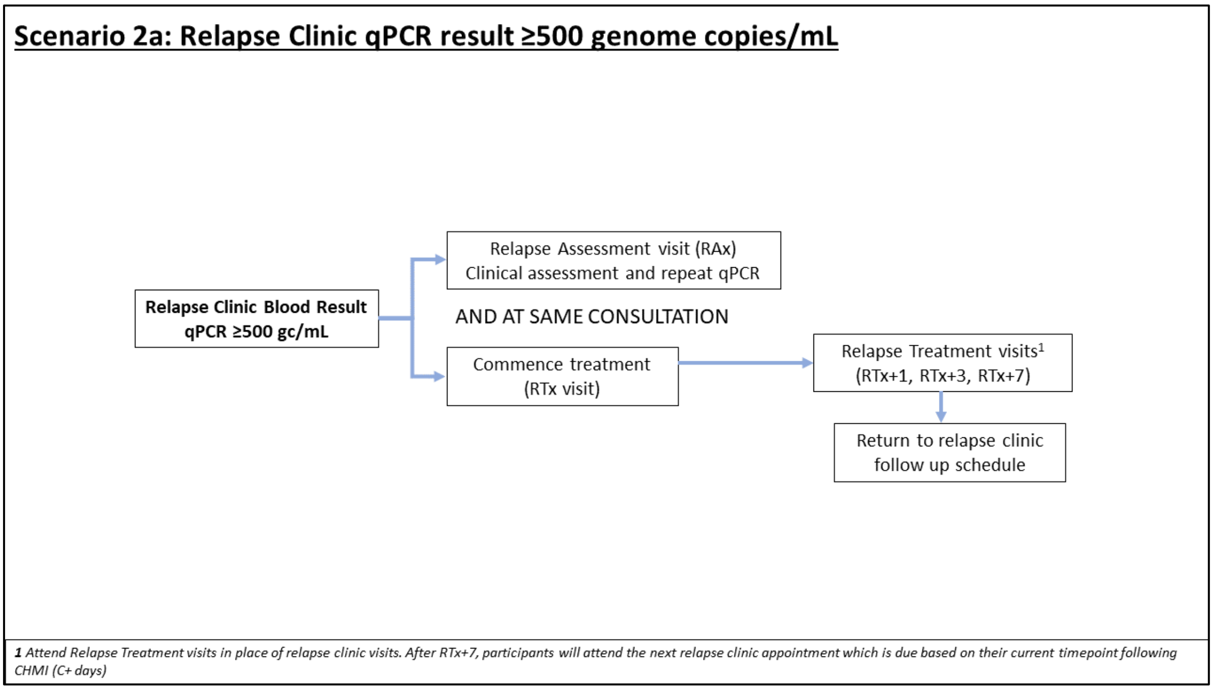

### Scenario 2b: Relapse Clinic qPCR result positive but <500 genome copies/mL

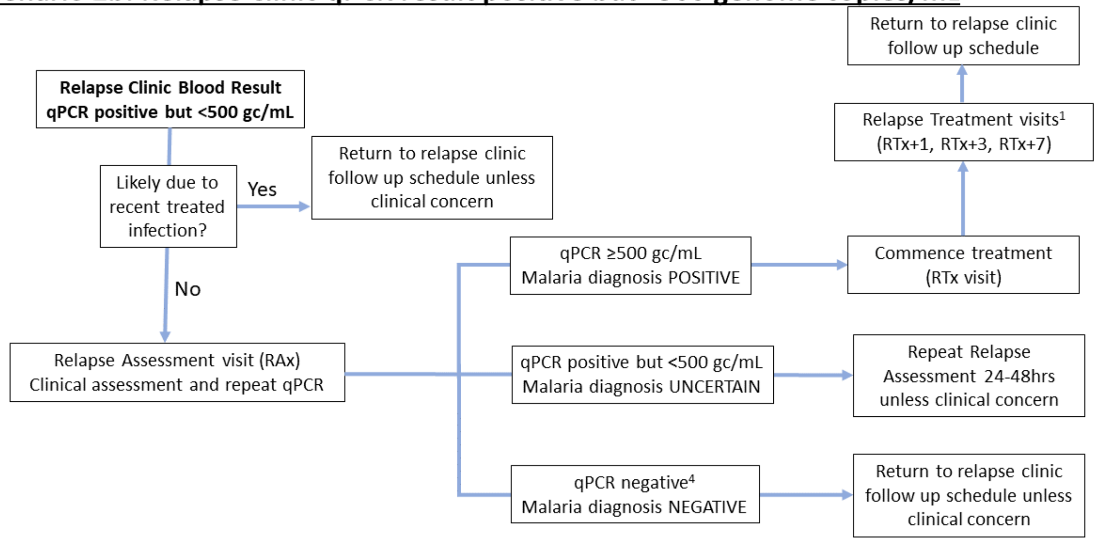

***1** Attend Relapse Treatment visits in place of relapse clinic visits. After RTx+7, participants will attend the next relapse clinic appointment which is due based on their current timepoint following CHMI (C+ days); **4** Negative as defined in qPCR SOP*

### Scenario 2c: Relapse Clinic qPCR result <20 genome copies/mL

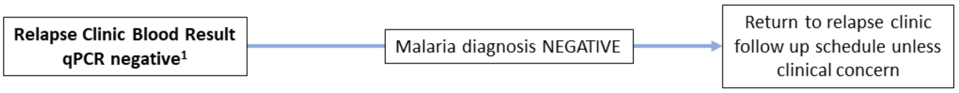

***1** Negative as defined in qPCR SOP*

#### Malaria parasite quantification by qPCR

DNA was extracted from 200 µL of whole blood using KingFisher Apex™ (ThermoFisher Scientific, 5400930P) and MagMAX DNA Multi-Sample Ultra 2.0 Kit (ThermoFisher Scientific, A36570). DNA was eluted in 100 µL of kit Elution Buffer and run in triplicate in qPCR assay as follows. 5 µL DNA sample was added to 20 µL of reaction mix consisting of 1x Luna qPCR Probe Universal Master Mix (New England Biolabs, M3004), primers at 0·4 µM and probe at 0·2 µM (Applied Biosystems).

Primers and probe target a pan-*Plasmodium* specific sequence in the 18S RNA gene (Forward: 5'-AGG AAG TTT AAG GCA ACA ACA GGT-3'; Reverse: 5'-GCA ATA ATC TAT CCC CAT CAC GA-3', Probe: 5'-FAM-TGA ACT AGG CTG CAC GCG-MGB-3'). Primers are as per McCarthy *et al.,* with probe sequence adapted to use FAM.[2] qPCR was conducted using a QuantStudio3 (ThermoFisher Scientific, A28132): with initial 95°C, 1 min step; followed by 40 cycles: 95°C for 15 s, 60°C for 30 s, with ROX passive reference.

Standards generated from a linearised plasmid were used to quantify parasites in samples,[2] and standards concorded to previously quantified samples from prior CHMI samples ahead of study start. Triplicates of each standard and non-template control wells were run in every assay. Standard curves and samples were analysed using Design & Analysis Software v2.8.0 9 (with Primary Analysis v1.8.1, Standard Curve v1.8.0 plugins) (ThermoFisher Scientific). Standard curves had to pass initial QC parameters of $R^2 \geq 0{\cdot}98$ and Efficiency 90-110%. The Lower Limit of Detection (LLOD) and Lower Limit of Quantification (LLOQ) were determined as 47 and 209 genome copies/mL, respectively.

#### Expression and purification of recombinant PvMSP1-19 protein

The gene sequence of PvMSP1-19 from strain Sal-1 was obtained from PlasmoDB, before synthesis by Twist Bioscience into a C-Tag expression vector. Recombinant protein was expressed in Expi293 cells using Expifectamine™ transfection kit (Thermo Fisher Scientific) as per manufacturer's instructions, before purification using C-tag affinity chromatography on the AKTA pure chromatography system (Cytiva), followed by SEC into TBS pH7·4 (20 mM Tris-HCl, 150 mM NaCl).

#### Anti-PvMSP1-19 enzyme-linked immunosorbent assay (ELISA)

Anti-PvMSP1-19 serum IgG responses were measured by endpoint enzyme-linked immunosorbent assay (ELISA). Briefly, 96-well flat bottom NUNC Maxisorp plates (Thermo Scientific) were coated overnight with PvMSP1-19 protein in Casein blocker in PBS (Casein, Thermo Scientific) at a concentration of 2 µg/mL at 4 °C. The plates were then washed with PBS/0·05% Tween-20 (PBS/T) before being blocked with Casein for 1 h at room temperature (RT). Test serum samples were then diluted 1 in 500 in Casein, added in duplicate to the plate and serially diluted 3-fold down the plate. A high-responding serially-diluted serum sample (positive control) and four casein-only (negative control) wells were included and incubated at RT for another 2 h. The plates were then washed again in PBS/T and Anti-Human IgG (γ-chain specific) Alkaline Phosphatase antibody (Sigma) was diluted 1 in 1000 in Casein and added at RT for 1 h. The plates were then washed for a final time and development buffer (*p*-nitrophenyl phosphate substrate (Sigma) diluted in diethanolamine buffer (Thermo

Scientific)) was added and optical density (OD) at 405nm ($OD_{405}$) read using an ELx808 microplate reader (BioTek, UK). Endpoint titres were calculated by determining the point at which the dilution curve intercepts the x axis at baseline fixed absorbance value of 0·15 $OD_{405}$.

### Supplementary Results

### CYP2D6 results

**Table S3: CYP2D6 results of enrolled participants (n=5)**

| | Participant | | | | |
|---|---|---|---|---|---|
| | **001** | **002** | **003** | **004** | **005** |
| **CYP2D6 phenotype** | Extensive metaboliser | Extensive metaboliser | Extensive metaboliser | Extensive metaboliser | Extensive metaboliser |
| **Predicted CYP2D6 genotype** | *1 / *1 | *2 / (*35 / *41) | *2 / (*2 / *41) | *1 / *1 | *2 / (*2 / *35) |
| **Activity score**[+] | 2.·0 | 1.·25–2·0 | 1.·25–2·0 | 2.·0 | 2.·0 |
| CYP2D6*2 (886C>T) | C/C Wild type | T/T Homozygous mutation | T/T Homozygous mutation | C/C Wild type | T/T Homozygous mutation |
| CYP2D6*2 (1457G>C) | G/G Wild type | C/C Homozygous mutation | C/C Homozygous mutation | G/G Wild type | C/C Homozygous mutation |
| CYP2D6*3 (2549delA) | A/A Wild type | A/A Wild type | A/A Wild type | A/A Wild type | A/A Wild type |
| CYP2D6*4 (1846G>A) | G/G Wild type | G/G Wild type | G/G Wild type | G/G Wild type | G/G Wild type |
| CYP2D6*5 (Gene Deletion) | Deletion not detected | Deletion not detected | Deletion not detected | Deletion not detected | Deletion not detected |
| CYP2D6*6 (1707delT) | T/T Wild type | T/T Wild type | T/T Wild type | T/T Wild type | T/T Wild type |
| CYP2D6*7 (2935A>C | A/A Wild type | A/A Wild type | A/A Wild type | A/A Wild type | A/A Wild type |
| CYP2D6*8 (1758G>T) | G/G Wild type | G/G Wild type | G/G Wild type | G/G Wild type | G/G Wild type |
| CYP2D6*9 (2615_2617delAAG) | AAG/AAG Wild type | AAG/AAG Wild type | AAG/AAG Wild type | AAG/AAG Wild type | AAG/AAG Wild type |
| CYP2D6*10 (100C>T) | C/C Wild type | C/C Wild type | C/C Wild type | C/C Wild type | C/C Wild type |
| CYP2D6*11 (883G>C) | G/G Wild type | G/G Wild type | G/G Wild type | G/G Wild type | G/G Wild type |
| CYP2D6*12 (124G>A) | G/G Wild type | G/G Wild type | G/G Wild type | G/G Wild type | G/G Wild type |
| CYP2D6*17 (1023C>T) | C/C Wild type | C/C Wild type | C/C Wild type | C/C Wild type | C/C Wild type |
| CYP2D6*29 (3183G>A) | G/G Wild type | G/G Wild type | G/G Wild type | G/G Wild type | G/G Wild type |
| CYP2D6*35 (31G>A) | G/G Wild type | G/A Heterozygous mutation | G/G Wild type | G/G Wild type | G/A Heterozygous mutation |
| CYP2D6*41 (2988G>A) | G/G Wild type | G/A Heterozygous mutation | G/A Heterozygous mutation | G/G Wild type | G/G Wild type |
| CYP2D6*43 (77G>A) | G/G Wild type | G/G Wild type | G/G Wild type | G/G Wild type | G/G Wild type |
| CYP2D6*45 (1717G>A) | G/G Wild type | G/G Wild type | G/G Wild type | G/G Wild type | G/G Wild type |
| CYP2D6*46 (77G>A) | G/G Wild type | G/G Wild type | G/G Wild type | G/G Wild type | G/G Wild type |
| CYP2D6*46 (1717G>A) | G/G Wild type | G/G Wild type | G/G Wild type | G/G Wild type | G/G Wild type |
| CYP2D6*xN (Gene Duplication) | Duplication not detected | Duplication not detected | Duplication not detected | Duplication not detected | Duplication not detected |

CYP2D6 = Cytochrome P450 2D6. CYP2D6 phenotype (poor, intermediate, extensive or ultra-rapid metaboliser) and activity score according to resources available at ClinPGx (clinpgx.org).[1] Extensive metaboliser phenotype activity score ≥ 1·25.

### Mosquito-bite CHMI

Participants 001, 002 and 003 received mosquitoes from Batch A which had been infected by DMFA with prior Percoll enrichment using blood from a RaViCHMI1 volunteer on Day 19 following blood-stage CHMI (after initiation of mepacrine treatment). Batch A mosquitoes had a mean of 49·1 oocysts per mosquito. A sample of Batch A mosquitoes (n=10) dissected one day prior to CHMI had a mean of 40,250 sporozoites per mosquito.

Participants 004 and 005 received mosquitoes from Batch B which had been infected by DMFA with prior Percoll enrichment using blood from a RaViCHMI1 volunteer on Day 19 following blood-stage CHMI (prior to any drug treatment). Batch B mosquitoes had a mean of 28·3 oocysts per mosquito. A sample of Batch B mosquitoes (n=10) dissected one day prior to CHMI had a mean of 46,500 sporozoites per mosquito.

Each participant (n=5) was exposed to five mosquitoes for 10 min after which the mosquitoes were individually dissected to confirm that i) the mosquito had taken a blood meal and ii) salivary glands were positive for sporozoites. On dissection, 25/25 mosquitoes had evidence of taking a blood meal and 24/25 had evidence of sporozoite positivity. The affected participant was exposed to one more mosquito for 10 min which subsequently fulfilled the above criteria on dissection.

Participants were diagnosed with primary *P. vivax* infection (qPCR > 500 genome copies/mL) 8–10 days following CHMI (figure S1).

**Figure S1: Kaplan-Meier plot of time to diagnosis (qPCR >500 genome copies/mL) of primary *P. vivax* infection**

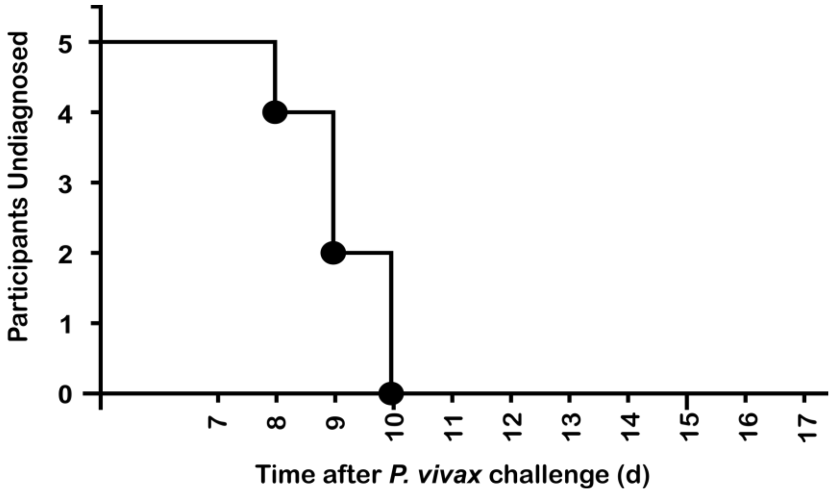

### Adverse events

**Figure S2: Solicited adverse events during primary and relapse *P. vivax* infections**

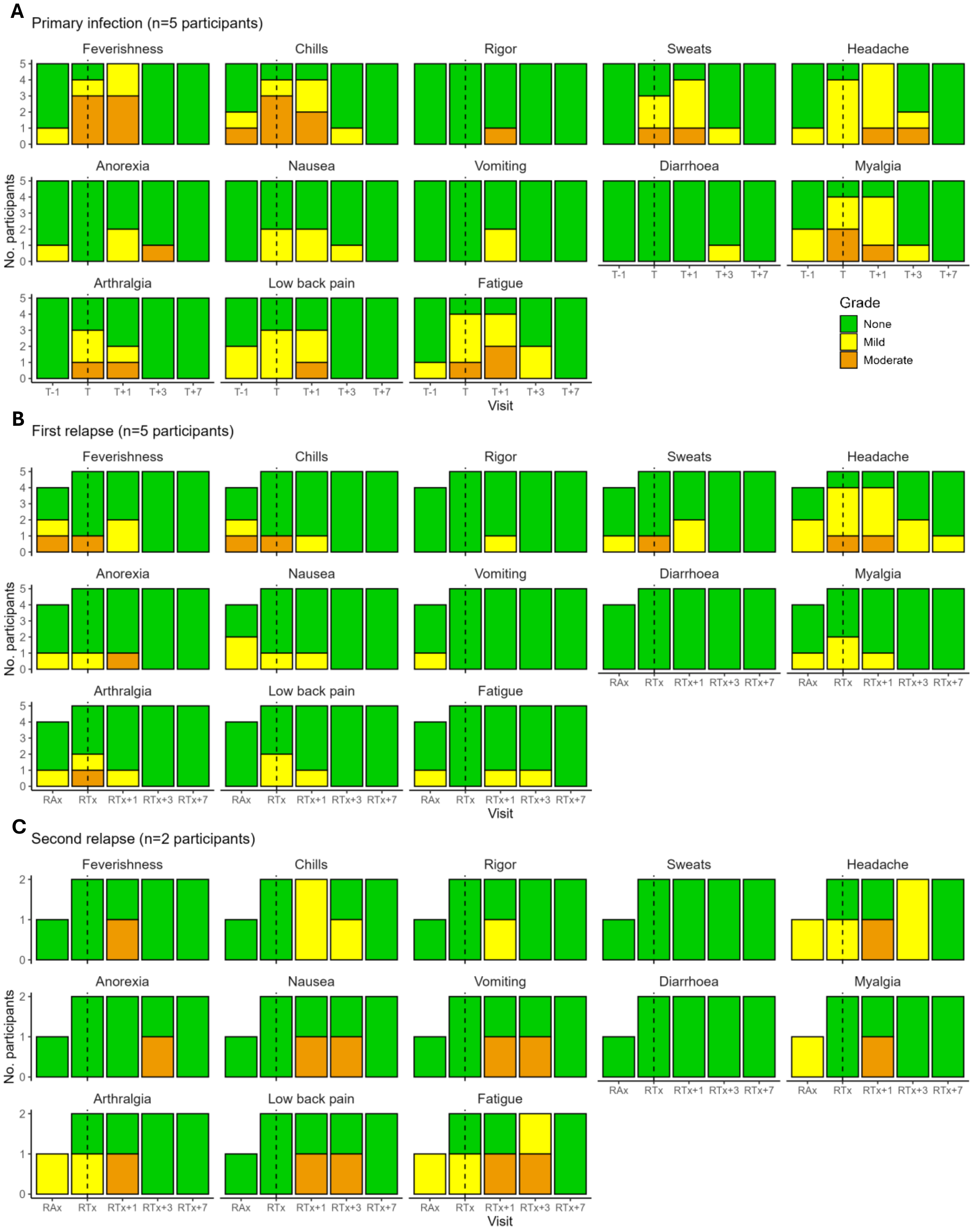

Number of participants reporting solicited adverse events at scheduled study visits during A) Primary, B) First relapse and C) Second relapse infections. A) T-1 = Day prior to treatment initiation; T = Day of treatment initiation; T+1, T+3, T+7 = 1, 3, 7 days after initiation of treatment respectively. B) and C) RAx = Relapse assessment visit; RTx = Day of treatment of relapse infection; RTx+1, RTx+3, RTx+7 = 1, 3, 7 days after initiation of treatment respectively. Vertical dashed line at treatment initiation of primary or relapse infection. Colour indicates severity grade. Two participants (one first relapse and one second relapse) did not attend RAx visit as they were diagnosed by qPCR at definitive treatment visit.

**Figure S3: Total number of solicited adverse events during primary and relapse *P. vivax* infections**

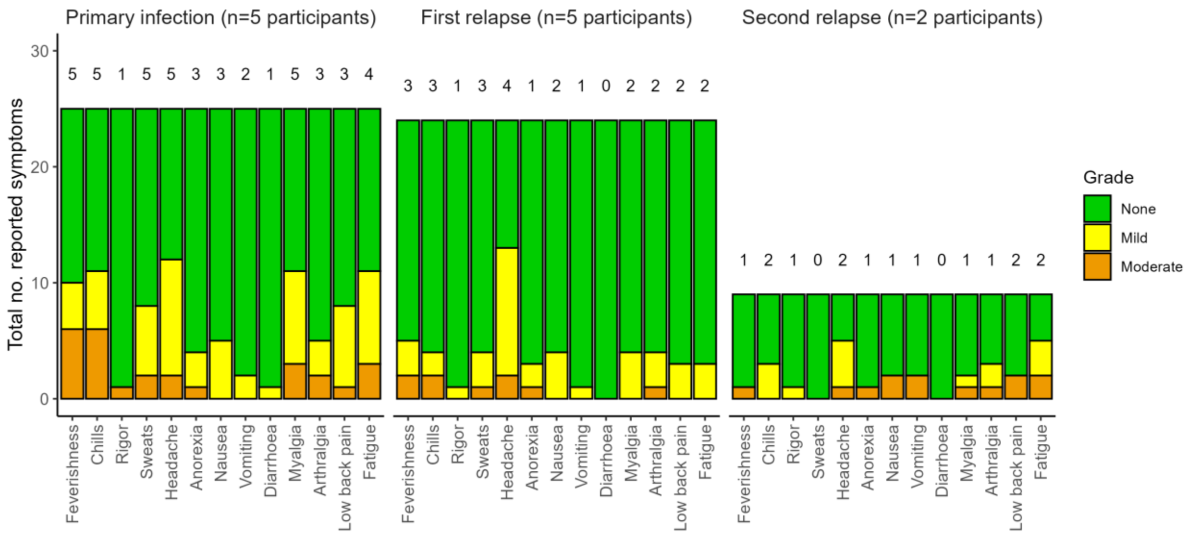

Total number of solicited adverse events reported by all participants at scheduled study visits during primary, first relapse and second relapse infections. Data included in primary infection are from T-1 = Day prior to treatment initiation; T = Day of treatment initiation; T+1, T+3, T+7 = 1, 3, 7 days after initiation of treatment respectively. Data included in relapse infections are from RAx = Relapse assessment visit; RTx = Day of treatment of relapse infection; RTx+1, RTx+3, RTx+7 = 1, 3, 7 days after initiation of treatment respectively. Number above bar indicates the number of participants reporting symptom at any severity (mild, moderate, severe). Colour indicates severity grade. Two participants (one first relapse and one second relapse) did not attend RAx visit as they were diagnosed by qPCR at definitive treatment visit.

**Figure S4: Maximum temperature recorded during primary and relapse *P. vivax* infections**

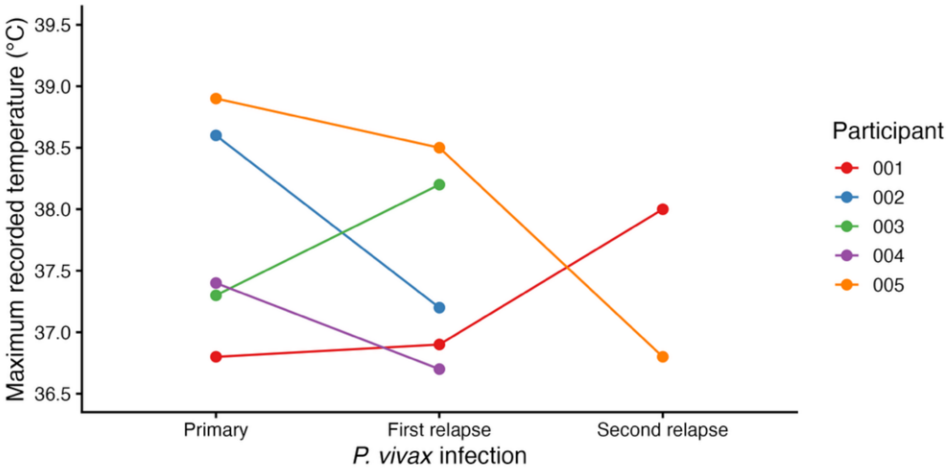

Maximum temperature recorded for each participant during primary, first relapse and second relapse *P. vivax* infections.

**Table S4: Solicited adverse events associated with mosquito bites (n=5)**

| | | Severity | | |
|---|---|---|---|---|
| System Order Class | Preferred Term | Grade 1 | Grade 2 | Grade 3 |
| Skin and subcutaneous tissue disorders | Erythema | 5 | | |
| | Pruritus | 5 | | |
| | Skin swelling | 3 | | |

Data are number of solicited adverse events associated with mosquito bites presented as maximum severity per episode.

**Table S5: Unsolicited adverse events associated with *P. vivax* infection (n=5)**

| | | Severity | | | ***P. vivax* infection and start of adverse event** |
|---|---|---|---|---|---|
| System Order Class | Preferred Term | Grade 1 | Grade 2 | Grade 3 | |
| Gastrointestinal disorders | Abdominal pain | | | 1* | Second relapse infection<br>2 days following diagnosis |
| Infections and infestations | Oral herpes | 1 | | | First relapse infection<br>1 day prior to diagnosis |
| Metabolism and nutrition disorders | Dehydration | | | 1* | Second relapse infection<br>2 days following diagnosis |

Data are number of unsolicited adverse events deemed at least possibly related to *P. vivax* infection (primary or relapse). Data presented as maximum severity per episode listed by MedDRA System Organ Class and Preferred Term. *Occurred in same participant and also deemed probably related to anti-malarial medication (atovaquone-proguanil).

**Table S6: Adverse events associated with anti-malarial medication**

| | | Severity | | | Stage of *P. vivax* treatment and start of adverse event | Medication |
|---|---|---|---|---|---|---|
| System Order Class | Preferred Term | Grade 1 | Grade 2 | Grade 3 | | |
| Blood and lymphatic system disorders | Methaemoglobinaemia | | 1+ | | Definitive malaria treatment (no active infection)<br><br>11 days following initiation of definitive treatment<br><br>Day 9 of primaquine treatment | Primaquine |
| Cardiac disorders | Palpitations | 1 | | | Primary infection<br><br>3 days following initiation of treatment | Artemether-lumefantrine |
| Gastrointestinal disorders | Abdominal pain | | | 1* | Second relapse infection<br><br>2 days following initiation of treatment | Atovaquone-proguanil |
| | Diarrhoea | | 1 | | Definitive malaria treatment (no active infection)<br><br>Same day as initiation of definitive treatment | Atovaquone-proguanil |
| | Nausea | 1+ | | | Definitive malaria treatment (no active infection)<br><br>3 days following initiation of definitive treatment<br><br>Day 1 of primaquine treatment | Primaquine |
| Metabolism and nutrition disorders | Dehydration | | | 1* | Second relapse infection<br><br>2 days following initiation of treatment | Atovaquone-proguanil |

Data are number of adverse events deemed at least possibly related to anti-malarial medication. Data presented as maximum severity per episode listed by MedDRA System Organ Class and Preferred Term. +Occurred in same participant. *Occurred in same participant and also deemed possibly related to second relapse *P. vivax* infection.

**Table S7: Laboratory adverse events**

| | | Primary infection (n=5) | | | First relapse infection (n=5) | | | Second relapse infection (n=2) | | |
|---|---|---|---|---|---|---|---|---|---|---|
| | | Grade 1 | Grade 2 | Grade 3 | Grade 1 | Grade 2 | Grade 3 | Grade 1 | Grade 2 | Grade 3 |
| Haematology | Anaemia | 3 | | | 3 | | 1 | 2 | | |
| | Leukopenia | 1 | | | 1 | | | | | |
| | Neutropenia | | | | 1 | 1 | | | | |
| | Lymphopenia | 1 | 3 | | 1 | 1 | | | | 1 |
| | Thrombocytopenia | | | 1 | | | 1 | 1 | | |
| Biochemistry | Hypokalaemia | | | 1 | | | | | | |
| | Elevated urea | | 1 | | | | | | | |
| | Elevated ALT | 2 | | | | | | | | |

Data are number of laboratory adverse events deemed at least possibly related to *P. vivax* infection (primary or relapse), presented as maximum severity per episode. ALT = Alanine transaminase.

**Figure S5: Lymphocyte and platelet count during primary and relapse *P. vivax* infections**

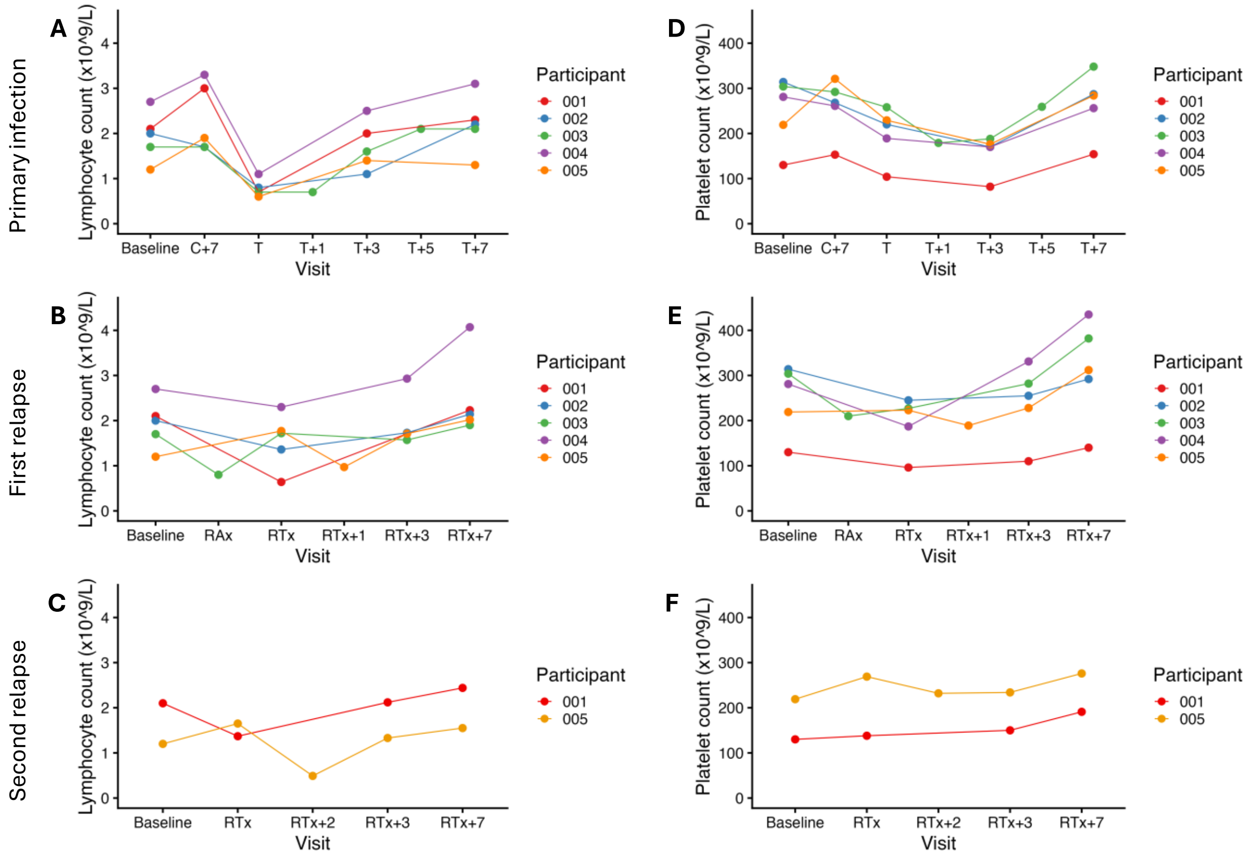

Lymphocyte count (x$10^9$/L) over time for each volunteer at A) primary B) first relapse and C) second relapse *P. vivax* infections. Platelet count (x$10^9$/L) over time for each volunteer at D) primary E) first relapse and F) second relapse *P. vivax* infection. Colour represents individual participant. The following timepoints are shown: Baseline = 2 days before CHMI; C+7 = 7 days after CHMI; T = Day of treatment of primary infection; T+1, T+3, T+5, T+7 = 1, 3, 5, 7 days after initiation of treatment respectively; RAx = Relapse assessment visit; RTx = Day of treatment of relapse infection; RTx+1, RTx+2, RTx+3, RTx+7 = 1, 2, 3, 7 days after initiation of treatment respectively. Blood sampling at T+1, T+5, RAx, RTx+1 and RTx+2 was performed at the discretion of the study physician. All other visits included blood sampling per protocol.

### Parasitaemia by qPCR

**Figure S6: Maximum detected *P. vivax* parasitaemia experienced by each participant during primary and relapse infections as measured by qPCR in genome copies/mL**

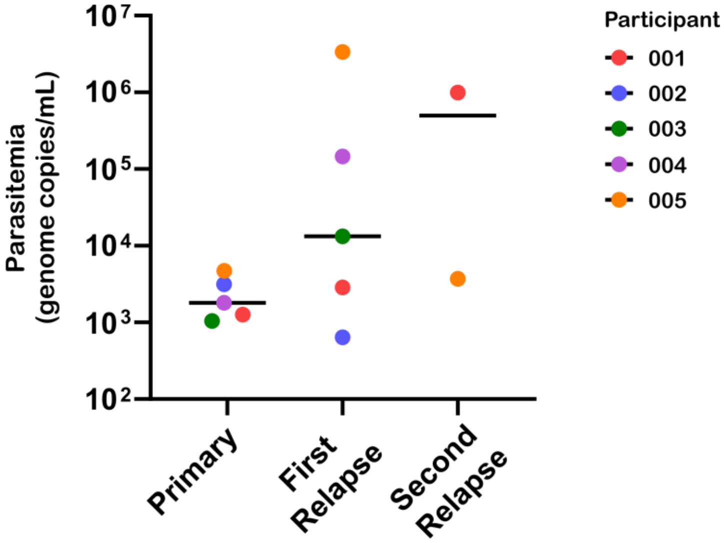

Maximum recorded parasitaemia for each participant measured by qPCR during primary, first relapse and second relapse *P. vivax* infections. Mean of triplicate values within an assay plotted for each participant. Bar at median. The difference between peak parasitaemia at primary infection (n=5) and all relapse infections (n=7) was not statistically significant (Wilcoxon signed-rank test, p = 0·15). The difference between peak parasitaemia at primary and first relapse infection (n=5, paired data) was not statistically significant (Wilcoxon signed-rank test for paired data, p = 0·19).

**Table S8: Triplicate parasitaemia values determined by qPCR in genome copies/mL during primary *P. vivax* infection for each participant.**

| | **Days after challenge** | **-2** | **7** | **8** | **9** | **10** | **11** | **12** | **13** | **14** | **15** | **16** | **17** |
|---|---|---|---|---|---|---|---|---|---|---|---|---|---|
| | **Trial Visit** | **C-2** | **C+7** | **C+8** | **C+9** | **C+10** | **C+11** | **C+12** | **C+13** | **C+14** | **C+15** | **C+16** | **C+17** |
| **Participant** | **001** | 0 | 0 | 36 | 606 | 1290 | 0 | n.d. | 0 | n.d. | n.d. | 0 | n.d. |
| | | 0 | 0 | 139 | 499 | 1442 | 0 | n.d. | 0 | n.d. | n.d. | 0 | n.d. |
| | | 0 | 0 | 57 | 418 | 1069 | 0 | n.d. | 0 | n.d. | n.d. | 0 | n.d. |
| | **002** | 0 | 0 | 1648 | 3013 | 0 | n.d. | 0 | n.d. | n.d. | n.d. | 0 | n.d. |
| | | 0 | 0 | 1710 | 3037 | 10 | n.d. | 0 | n.d. | n.d. | n.d. | 0 | n.d. |
| | | 0 | 0 | 2044 | 3381 | 23 | n.d. | 0 | n.d. | n.d. | n.d. | 0 | n.d. |
| | **003** | 0 | 0 | 31 | 778 | 1087 | 0 | n.d. | 0 | n.d. | n.d. | n.d. | 0 |
| | | 0 | 0 | 60 | 615 | 996 | 0 | n.d. | 0 | n.d. | n.d. | n.d. | 0 |
| | | 0 | 0 | 66 | 532 | 1043 | 0 | n.d. | 0 | n.d. | n.d. | n.d. | 0 |
| | **004** | 0 | 0 | 131 | 257 | 824 | 1665 | 0 | n.d. | 0 | n.d. | n.d. | 0 |
| | | 0 | 0 | 62 | 164 | 584 | 1901 | 0 | n.d. | 0 | n.d. | n.d. | 0 |
| | | 0 | 0 | 32 | 186 | 850 | 1823 | 0 | n.d. | 0 | n.d. | n.d. | 0 |
| | **005** | 0 | 0 | 131 | *217* | 1317 | 5496 | 0 | n.d. | 0 | n.d. | n.d. | 0 |
| | | 0 | 0 | 36 | *445* | 1280 | 4608 | 0 | n.d. | 0 | n.d. | n.d. | 0 |
| | | 0 | 0 | 153 | *267* | 1017 | 4054 | 0 | n.d. | 0 | n.d. | n.d. | 0 |

n.d. = no data. C+X = timepoint associated with scheduled clinic visit X days following CHMI. Green: Day of reaching threshold for treatment (qPCR >500 genome copies/mL); Blue: T = Day of treatment initiation; Orange: T+1, T+3, T+7 = 1, 3, 7 days after initiation of treatment respectively. LLOD and LLOQ determined as 47 and 209 genome copies/mL, respectively. Italics: data point included after passing secondary QC (see supplementary methods).

**Table S9: Triplicate parasitaemia values determined by qPCR in genome copies/mL during relapse follow-up period for each participant**.

**A. Scheduled fortnightly clinics for each participant**

| | **Days after challenge** | **29** | **43** | **57** | **71/72** | **85** | **99** | **113** | **115** | **127** | **141** | **155** | **169** | **183** |
|---|---|---|---|---|---|---|---|---|---|---|---|---|---|---|
| | **Trial Visit** | **C+28** | **C+42** | **C+56** | **C+70** | **C+84** | **C+98** | **C+112** | **C+112** | **C+126** | **C+140** | **C+154** | **C+168** | **C+182** |
| **Participant** | **001** | 0 | 0 | 0 | 0 | 0 | *122* | n.d. | 0 | 0 | 0 | 0 | 0 | 1016266 |
| | | 0 | 0 | 0 | 0 | 0 | *189* | n.d. | 0 | 0 | 0 | 0 | 0 | 1013789 |
| | | 0 | 0 | 0 | 0 | 0 | *72* | n.d. | 0 | 0 | 0 | 0 | 0 | 958216 |
| | **002** | 0 | 0 | 0 | 0 | 0 | 87 | 0 | n.d. | 0 | 0 | n.d. | n.d. | n.d. |
| | | 0 | 0 | 0 | 0 | 0 | 109 | 0 | n.d. | 0 | 0 | n.d. | n.d. | n.d. |
| | | 0 | 0 | 0 | 0 | 0 | 112 | 0 | n.d. | 0 | 0 | n.d. | n.d. | n.d. |
| | **003** | 0 | n.d.* | 0 | n.d.** | n.d. | n.d. | p.w. | p.w. | p.w. | p.w. | p.w. | p.w. | p.w. |
| | | 0 | n.d. | 0 | n.d. | n.d. | n.d. | p.w. | p.w. | p.w. | p.w. | p.w. | p.w. | p.w. |
| | | 0 | n.d. | 0 | n.d. | n.d. | n.d. | p.w. | p.w. | p.w. | p.w. | p.w. | p.w. | p.w. |
| | **004** | 0 | 0 | 0 | 0 | 0 | *44* | 0 | n.d. | 0 | 0 | 0 | 0 | 0 |
| | | 0 | 0 | 0 | 0 | 0 | *14* | 0 | n.d. | 0 | 0 | 0 | 0 | 0 |
| | | 0 | 0 | 0 | 0 | 0 | *25* | 0 | n.d. | 0 | 0 | 0 | 0 | 0 |
| | **005** | 0 | 0 | 0 | 0 | 0 | 0 | 0 | n.d. | 0 | 0 | 3241964 | 0 | 0 |
| | | 0 | 0 | 0 | 0 | 0 | 0 | 0 | n.d. | 0 | 0 | 3655547 | 0 | 0 |
| | | 0 | 0 | 0 | 0 | 0 | 0 | 0 | n.d. | 0 | 0 | 2877430 | 0 | 0 |

n.d. = no data. p.w. = participant withdrawn. C+X = timepoint associated with scheduled fortnightly clinic visit, X days following CHMI. Green: Day of reaching threshold for treatment (qPCR >500 genome copies/mL). LLOD and LLOQ determined as 47 and 209 genome copies/mL, respectively. Italics: data point included after passing secondary QC (see supplementary methods).
*C+42 visit coincided with RTx+7 for participant 003.
**in order to withdraw from study, participant 003 initiated definitive treatment on same day as C+70 clinic for other participants.

**B. Additional relapse assessments and any resultant relapse treatment and post-treatment timepoints for each participant**

| Participant | Relapse Assessment Event | Relapse Associated Timepoint | | | | | Confirmed Relapse (for Participant) |
|---|---|---|---|---|---|---|---|
| | | RAx | RTx | RTx+1 | RTx+3 | RTx+7 | |
| 001 | #1 | 1763<br>1710<br>1596 | 2814<br>2657<br>3067 | 0<br>0<br>0 | 0<br>0<br>0 | 0<br>0<br>0 | First |
| | #2 | C+182 visit* | 656515<br>630192<br>598987 | 41237<br>42225<br>40163 | 70<br>0<br>56 | 0<br>0<br>0 | Second |
| 002 | #1 | 0<br>0<br>0 | No relapse | | | | |
| | #2 | 122<br>169<br>173 | Initiated RAx3 | | | | |
| | #3 | 640<br>690<br>586 | 592<br>593<br>525 | 0<br>0<br>4 | 0<br>0<br>0 | 0<br>0<br>0 | First |
| 003 | #1 | 5407<br>6668<br>6319 | 14438<br>13739<br>11692 | 445<br>541<br>432 | 0<br>0<br>0 | 0<br>0<br>0 | First |
| 004 | #1 | 0<br>0<br>22 | Initiated RAx2 | | | | |
| | #2 | 0<br>0<br>0 | Initiated RAx3 | | | | |
| | #3 | 0<br>0<br>0 | No relapse | | | | |
| 005 | #1 | 0<br>0<br>0 | No relapse | | | | |
| | #2 | C+154 visit** | 3288214<br>3425568<br>3359687 | 394757<br>382933<br>360737 | 54443<br>63536<br>64208 | 0<br>0<br>0 | First |

RAx = Relapse assessment visit. RTx = Day of treatment of relapse infection. RTx+1, RTx+3, RTx+7 = 1, 3, 7 days after initiation of treatment respectively. Green: Day of reaching threshold for treatment (qPCR >500 genome copies/mL); Blue: Day treatment initiated (RTx or DDoT); Orange: Post-treatment samples. LLOD and LLOQ determined as 47 and 209 genome copies/mL, respectively. *Conducted 183 days following CHMI. **Conducted 155 days following CHMI.

**C. Definitive treatment timepoints for each participant**

| | | DDoT | DDoT+1 | DDoT+3 | DDoT+7 | DDoT+30 | Confirmed Relapse (for Participant) |
|---|---|---|---|---|---|---|---|
| Participant | 001 | 0<br>0<br>0 | 0<br>0<br>0 | 0<br>0<br>0 | 0<br>0<br>0 | 0<br>0<br>0 | |
| | 002 | 0<br>18<br>0 | 0<br>0<br>0 | 0<br>0<br>0 | 0<br>0<br>0 | 0<br>0<br>0 | |
| | 003 | 0<br>0<br>0 | 0<br>0<br>0 | 0<br>0<br>0 | 0<br>0<br>0 | 0<br>0<br>0 | |
| | 004 | 149829<br>147089<br>143113 | 11670<br>11468<br>12656 | 405<br>312<br>294 | 0<br>0<br>0 | 0<br>0<br>0 | First |
| | 005 | 3197<br>4355<br>3611 | 1533<br>1525<br>1822 | 670<br>756<br>805 | 0<br>0<br>0 | 0<br>0<br>0 | Second |

DDoT = Day of definitive treatment initiation. DDoT+1, DDoT+3, DDoT+7, DDoT+30 = 1, 3, 7, 30 days after definitive treatment initiation respectively. Blue: Day treatment initiated (RTx or DDoT); Orange: Post-treatment samples. LLOD and LLOQ determined as 47 and 209 genome copies/mL, respectively.

### Further details regarding presentation of *P. vivax* relapse infections

**Table S10: Further details regarding presentation of relapse *P. vivax* infections**

| Participant | Days following CHMI | Confirmed relapse | Maximum detected parasitaemia (genome copies/mL) | Anti-malarial | Presentation |
|---|---|---|---|---|---|
| **003** | 36 | First relapse | 13,289 | Artemether-lumefantrine | Participant presented with moderate feverishness, moderate chills, mild myalgia, mild fatigue and fever (38·2°C) on the evening of 12th day between fortnightly clinic visits. They were assessed the following morning and had a qPCR > 500 genome copies/mL (Day 36 following CHMI). Treatment with artemether-lumefantrine was initiated the following day. During the week prior to *P. vivax* relapse, they had experienced i) symptoms of a mild upper respiratory tract infection (cough and sore throat from Day 30 following CHMI) which had improved prior to relapse diagnosis, and ii) a cold sore (from Day 35 following CHMI). |
| **001** | 101 | First relapse | 2,846 | Artemether-lumefantrine | Participant had detectable parasitaemia on fortnightly qPCR in the absence of symptoms. This was initially below the threshold for treatment. They were reassessed at 48 hours (asymptomatic) at which point qPCR > 500 genome copies/mL (Day 101 following CHMI). Treatment with artemether-lumefantrine was initiated the following day. |
| **002** | 101 | First relapse | 639 | Artemether-lumefantrine | Participant had detectable parasitaemia on fortnightly qPCR in the absence of symptoms. This was initially below the threshold for treatment. They were reassessed at 24 hours (mild nausea and mild headache) and qPCR remained below threshold for treatment. They were reassessed after a further 24 hours (mild nausea and mild headache) at which point qPCR > 500 genome copies/mL (Day 101 following CHMI). Treatment with artemether-lumefantrine was initiated the following day. |
| **005** | 155 | First relapse | 3,357,823 | Artemether-lumefantrine | Participant presented to fortnightly clinic with moderate feverishness, moderate chills, mild sweats, mild headache, mild anorexia, mild nausea, mild vomiting and mild fatigue. qPCR > 500 genome copies/mL (Day 155 following CHMI). Treatment with artemether-lumefantrine was initiated the same day. |
| **001** | 183 | Second relapse | 996,090 | Artemether-lumefantrine | Participant presented to fortnightly clinic with mild headache, mild myalgia and mild arthralgia. qPCR > 500 genome copies/mL (Day 183 following CHMI). Treatment with artemether-lumefantrine was initiated the following day. During the week prior to *P. vivax* relapse, they had experienced symptoms of a mild upper respiratory tract infection (cough, nasal congestion, headache, sore throat and feverishness from Day 176 following CHMI) which had improved prior to relapse diagnosis. |
| **004** | 206 | First relapse | 146,677 | Atovaquone-proguanil | Participant attended clinic visit to commence definitive treatment (atovaquone-proguanil followed by primaquine). They did not report any symptoms. qPCR > 500 genome copies/mL. During the week prior to *P. vivax* relapse, they had experienced symptoms of a mild upper respiratory tract infection (cough, nasal congestion and headache from Day 201 following CHMI) which had improved prior to relapse diagnosis. |
| **005** | 206 | Second relapse | 3,721 | Atovaquone-proguanil | Participant attended clinic visit to commence definitive treatment (atovaquone-proguanil followed by primaquine). They did not report any symptoms. qPCR > 500 genome copies/mL. |

CHMI = Controlled Human Malaria Infection; qPCR = quantitative polymerase chain reaction.

### Climate during study period

#### Figure S7: Rainfall and temperature during study period

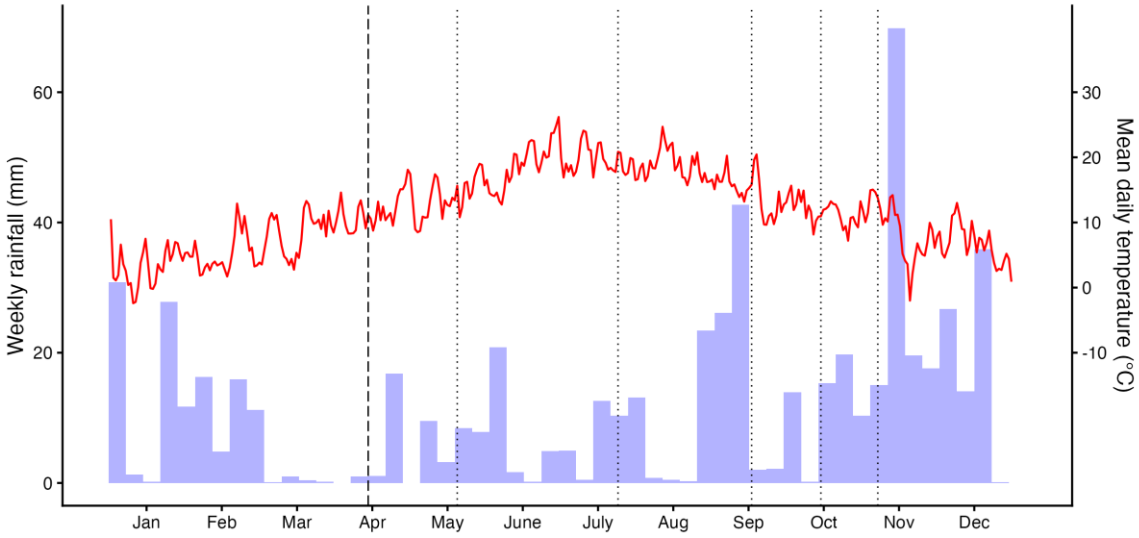

Weekly rainfall (blue bars) and mean daily temperature (red line) in Oxford, UK, 2025. Data from daily readings from the Radcliffe Meteorological Station, Green Templeton College, Oxford. Vertical dashed line indicates date of CHMI. Vertical dotted lines indicate diagnosis of relapse *P. vivax* infections. Relapses diagnosed at 36 days, 101 days (two participants), 155 days, 183 days, and 206 days (two participants) following CHMI.
